# Screening for Pathogenic Variants in Cardiomyopathy Genes Predicts Mortality and Composite Outcomes in UK Biobank

**DOI:** 10.1101/2022.06.27.22276949

**Authors:** Babken Asatryan, Ravi A. Shah, Ghaith Sharaf Dabbagh, Andrew P. Landstrom, Dawood Darbar, Mohammed Y Khanji, Luis R. Lopes, Stefan van Duijvenboden, Daniele Muser, Aaron Mark Lee, Christopher M. Haggerty, Pankaj Arora, Christopher Semsarian, Tobias Reichlin, Virend K. Somers, Anjali T. Owens, Steffen E. Petersen, Rajat Deo, Patricia B Munroe, Nay Aung, C. Anwar A. Chahal, the Genotype-First Approach Investigators

## Abstract

**Background:** Inherited cardiomyopathies can present with broad variation of phenotype. Data are limited regarding genetic screening strategies and outcomes associated with putative pathogenic variants (PuPV) in cardiomyopathy-associated genes in the general population.

**Objective:** We aimed to determine the risk of mortality and cardiomyopathy-related outcomes associated with PuPV in cardiomyopathy-associated genes in UK Biobank.

**Methods:** Using whole exome sequencing data, variants in dilated, hypertrophic and arrhythmogenic cardiomyopathy-associated genes with at least limited evidence of disease causality according to ClinGen Expert Panel curations, were annotated using REVEL (≥0.65) and ANNOVAR (predicted loss of function) to identify PuPVs. Individuals with PuPV comprised the genotype-positive (G+) and those without PuPV the genotype-negative (G-) cohorts. Group comparisons were made using time-to-event analyses for the primary (all-cause mortality) and secondary outcomes (diagnosis of cardiomyopathy; composite outcome of diagnosis of cardiomyopathy, heart failure, arrhythmia, stroke, and death).

**Results:** Among 200,619 participants, 22,401 (11.2%) were found to host ≥1 PuPV in cardiomyopathy-associated genes (G+). After adjusting for age and sex, G+ individuals had increased all-cause mortality [HR 1.07 (95%CI 1.02-1.13; p=0.011)] and increased rates of diagnosis of cardiomyopathy later in life [HR 2.37 (95%CI 1.98-2.85; p<0.0001)], which further increased in those with PuPV in definitive/strong evidence ClinGen genes [3.25 (95%CI 2.63-4.00; p<0.0001)]. G+ individuals had a higher risk of developing the composite outcome [HR 1.11 (95%CI 1.06-1.15; p<0.0001)].

**Conclusions:** Adults with PuPV in cardiomyopathy-associated genes have higher all-cause mortality and increased risk of developing cardiomyopathy-associated features and complications, compared to genotype-negative controls.

**Condensed Abstract:** Leveraging the UK Biobank prospective cohort, we analyzed whole exome sequencing data in dilated, hypertrophic and arrhythmogenic cardiomyopathy-associated genes using a population screening ‘genotype-first’ approach. Individuals with putative pathogenic variants in genes implicated in cardiomyopathies showed an increased risk of all-cause mortality, higher risk of developing clinical cardiomyopathy later in life, and higher risk of a composite outcome (cardiomyopathy, heart failure, arrhythmia, stroke, and death) compared to genotype-negative controls. These findings highlight the potential role of ‘genotype-first’ approach in elevating personalized medicine into population level precision health in the future.

## Introduction

Inherited cardiomyopathies (CMP), such as dilated (DCM), hypertrophic (HCM), and arrhythmogenic right ventricular cardiomyopathies (ARVC), are primarily genetic diseases associated with increased morbidity and mortality.(1) The majority of CMPs are autosomal dominant diseases with complex genetic architecture and demonstrate phenotypic heterogeneity, incomplete penetrance and variable expressivity, ranging from asymptomatic course to progressive disease with heart failure (HF), arrhythmias and stroke.(2) The sentinel event can be sudden cardiac death, underscoring the importance of early screening as a pathway to save lives and decrease morbidity.(3) Over 300 genes have been reported in association with inherited CMPs; however, a recent systematic evaluation of the evidence underlying these gene-disease associations by the Clinical Genome Resource (ClinGen) Gene Curation Expert Panels (GCEPs) found at least limited evidence for 44 genes in human monogenic DCM, 27 in HCM, and 18 in ARVC.(4-6)

Identifying individuals at risk for CMP-associated complications remains a challenge despite the enormous progress in diagnostic modalities, as a significant proportion of affected patients are diagnosed postmortem.(7) Underlying genotype is increasingly recognized as an important determinant of disease-related outcomes in clinically diagnosed patients.(2,8) With the decreasing cost of whole exome and whole genome sequencing (WES/WGS), a ‘genotype-first’ approach whereby individuals are evaluated based on their genetic variation, irrespective of phenotype, is emerging as a potentially powerful tool to solve such issues.(9) Despite many challenges, this approach can reveal a wide spectrum of phenotypic features and individuals with ‘concealed’ disease. Nonetheless, studies suggest that the clinical disease penetrance is low in putative pathogenic gene variant (PuPV) carriers at the population level.(10) However, data regarding whether CMP genotype can predict mortality, diagnosis of CMP later in life, or CMP-related outcomes are limited in the general population setting.(11) Thus, we leveraged the UK Biobank (UKBB) using a ‘genotype-first’ approach to test the hypotheses that individuals with PuPV in genes implicated in CMPs have increased mortality, higher risk of being diagnosed with cardiomyopathy and clinically relevant, CMP-related outcomes compared to genotype-negative controls.

## Methods

### Study population

The UKBB study is a prospective cohort study of 502,493 UK residents enrolled when aged between 40-69 years, recruited at 22 assessment centers across the UK.(12) Enrolled participants underwent comprehensive phenotyping at the respective assessment center. This included anthropometric measurements, answering extensive health and lifestyle questionnaires, and providing biological samples. This provided information on baseline characteristics and self-reported medical conditions. The UKBB additionally links to primary care records, and external hospital data records, providing, in the form of ICD-10 diagnostic and OPCS-4 operation codes, data from hospital admissions. Access to national death registries provides information on date and cause of death. Data were updated until February 2021, generating long-term follow-up data. A subset of participants in the UKBB have undergone WES. The UKBB received approval from the North West Multi-Centre Research Ethics Committee on June 17^th^, 2011 (Ref 11/NW/0382), which was extended on May 10^th^, 2016 (Ref 16/NW/0274) and extended on 18 June 2021 until 2026 (Ref 21/NW/0157) with written informed consent obtained from all participants.

### Selection of genes

The study cohort was formed by UKBB participants that had undergone WES. Briefly, exomes were captured with the IDT xGen Exome Research Panel v1.0 including supplemental probes. Primary and secondary analyses of the raw reads were performed using the Original Quality Functionally Equivalent pipeline.(13) Tertiary analyses were performed by annotating variants in a panel of 78 genes implicated in HCM, DCM, and/or ARVC by the respective ClinGen GCEPs.(4-6) A further group of genes considered to be causative for arrhythmogenic cardiomyopathy (ACM) (including right- or left-dominant forms) in the 2019 Heart Rhythm Society Expert Consensus Statement was selected for additional analysis.(14) Another subgroup of CMP genes, listed among the 78 actionable genes recommended by the American Council of Medical genetics and Genomics (ACMG) for reporting of secondary findings in clinical exome/genome sequencing, were also analyzed.(15) Details regarding gene selection, gene-disease associations and disease-causality evidence levels according to the ClinGen GCEPs are provided in the **Supplemental Methods**.

### Variant annotation pipeline

Our variant annotation pipeline was applied to this dataset and to the genes of interest, as published previously.(16) In brief, tertiary bioinformatic analysis was restricted to high quality and rare variants, which was defined as read depth ≥10, call quality ≥20, genotype quality ≥20, and minor allele frequency ≤0.001 in both gnomAD (17) and the UKBB exome dataset. ANNOVAR (18) and REVEL score (a method for predicting deleterious missense variants (19)) annotations were used to determine a set of PuPV, as used elsewhere.(20,21) Variants with ANNOVAR annotations of either frameshift insertions/deletions, gain/loss of stop codon, or disruption of canonical splice site dinucleotides were annotated as predicted loss-of-function variants (pLOF). Missense variants were annotated as predicted pathogenic missense variants if the annotated REVEL score was ≥0.65.(20) For *TTN*, only radical variants (i.e., nonsense, frameshift, and splice-site variants) were considered for analysis. A further disease specific filtering allele frequency (FAF) was applied, removing all variants with frequency more than the applied FAF in gnomAD and/or UKBB to produce our final set of PuPV variants.(22) The applied FAF was 8.4 × 10^−5^ for DCM associated variants, 4 × 10^−5^ for HCM associated variants, and 9.2 × 10^−5^ for ARVC associated variants.(22) Due to the population prevalence of these CMPs, variants that occur more frequent than these FAF are unlikely to be causative variants. These frequency thresholds have been previously defined.(16,22) Variants annotated as predicted pathogenic missense or pLOF at a frequency lower than the FAF formed the set of PuPV. Individuals with a PuPV comprised the genotype-positive (G+) cohort and those without formed the referent-control/genotype-negative (G-) cohort. Quality control analysis of our variant annotation strategy has been published recently.(16)

Phenotypic definitions were based on a combination of clinical diagnoses (self-reported conditions and ICD-10 codes) and procedures (self-reported and OPCS-4 codes). A full list of phenotype definitions, adapted from definitions used elsewhere,(23-25) is shown in **Table S1**. Clinical DCM was defined by the presence of ICD-10 code I42.0 or on CMR (LV end-diastolic volume >2SD from Z score corrected for body surface area and systolic dysfunction by decreased LV ejection fraction). Clinical HCM was defined by the presence of ICD-10 code I42.1, I42.2 or thickness of interventricular septum ≥15 mm on CMR. Clinical ARVC was defined by ICD-10 code I42.8 only.

Access to the UKBB was provided under application number 48286. UKBB participants who withdrew consent were excluded.

### Statistical analysis

Data were analyzed using *R 4*.*0* packages *tidyverse, survival*, and *tableone*. Continuous, normally distributed variables were summarized using mean and standard deviation and compared using two-sample *t*-tests. Non-normally distributed continuous data were summarized using median and interquartile range and compared using Wilcoxon-rank sum tests. Categorical data were summarized using percentages and compared using Chi-squared or Fisher’s exact tests.

Time-to-event analysis was performed using Cox proportional hazard regression. Survival duration was calculated from date of enrollment to date of censoring. Data was treated as left truncated (by date of enrollment) and right censored. Age was chosen as the timescale rather than time-on-study as this approach reduces bias associated with potential confounding by age in cohort studies.(26,27) The primary outcome was all-cause mortality. The secondary outcomes were 1) developing clinical cardiomyopathy later in life, and 2) composite outcome of any one of the following: HF, clinical diagnosis of CMP, stroke, cardiac implantable electronic device (CIED) implantation, atrial fibrillation, sustained and non-sustained ventricular tachyarrhythmia, and/or mortality. The proportional hazard assumption was tested by visual inspection of the scaled Schoenfeld residual versus transformed time plot and statistically by a Chi-squared test of scaled Schoenfeld residual and transformed time. Hazard ratios (HR) are reported with 95% confidence intervals (CI). This analysis was performed using the *survival* package. Both outcomes were adjusted for age and sex.

## Results

### Baseline characteristics

Of 200,619 UKBB participants with WES, 22,401 (11.2%) had ≥1 PuPV in CMP-associated genes (G+ cohort); this included 16,798 (8.4%) individuals with DCM-associated PuPV, 12,745 (6.4%) individuals with HCM-associated PuPV, and 8,028 (4.0%) individuals with ARVC-associated PuPV. G+ and G- groups had similar proportion of females (54.9% vs. 55.1%, respectively; p=0.645). Across all CMPs, G+ participants were slightly younger at enrollment (for all CMPs together: 56.2 vs 56.5 years; p<0.0001). Overall, 93.9% of the participants were White Caucasians (self-reported). Both in the overall CMP group, and in individual CMP genotype-based subgroups (DCM, HCM, ARVC), G+ individuals were more likely to receive the clinical diagnosis of the respective gene-associated and other CMPs, than the G- individuals, with the exception of those with ARVC-G+, who did not have higher rates of diagnosis of HCM (**Table 1, Table S1)**.

**Table 1.**
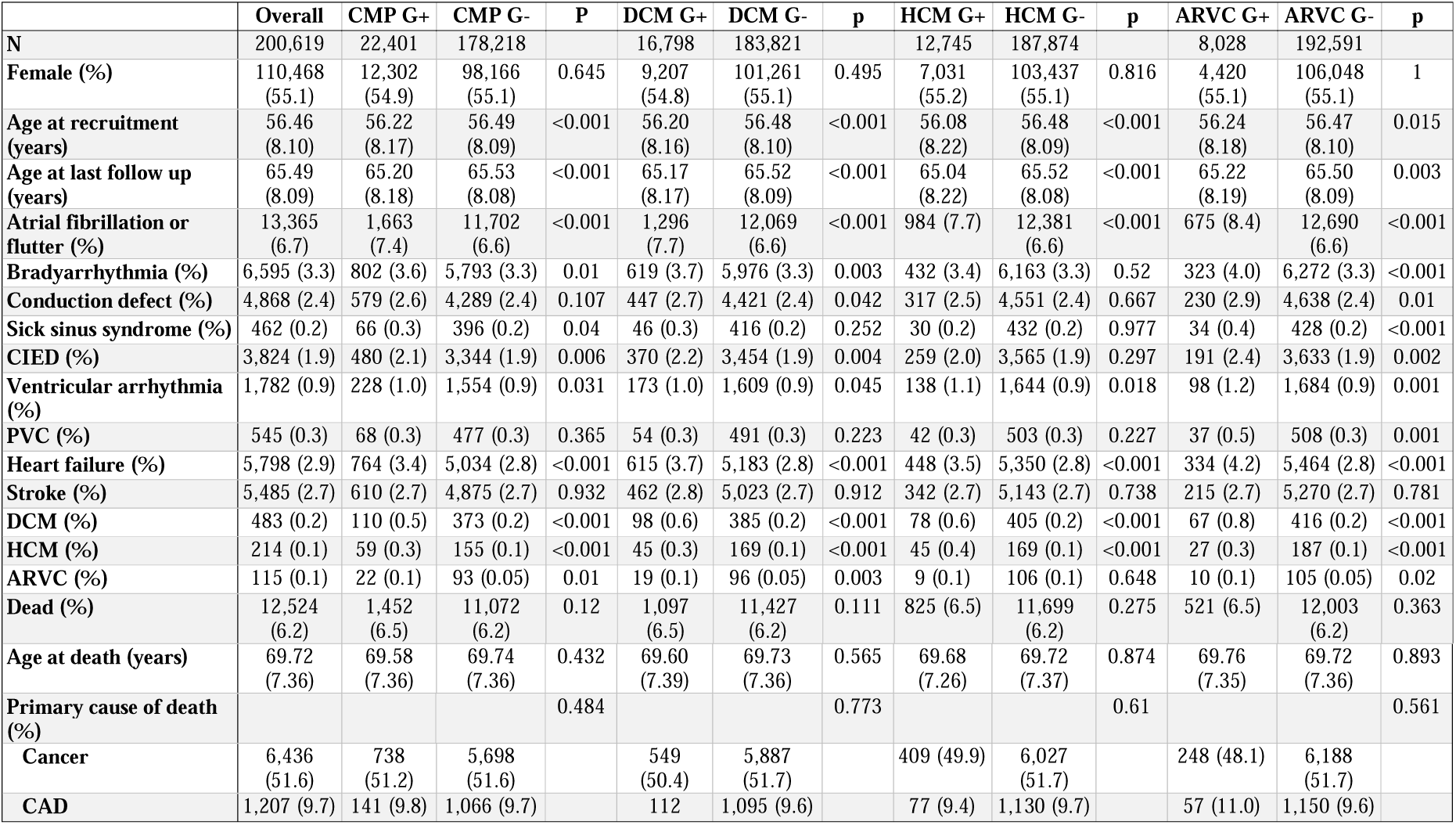
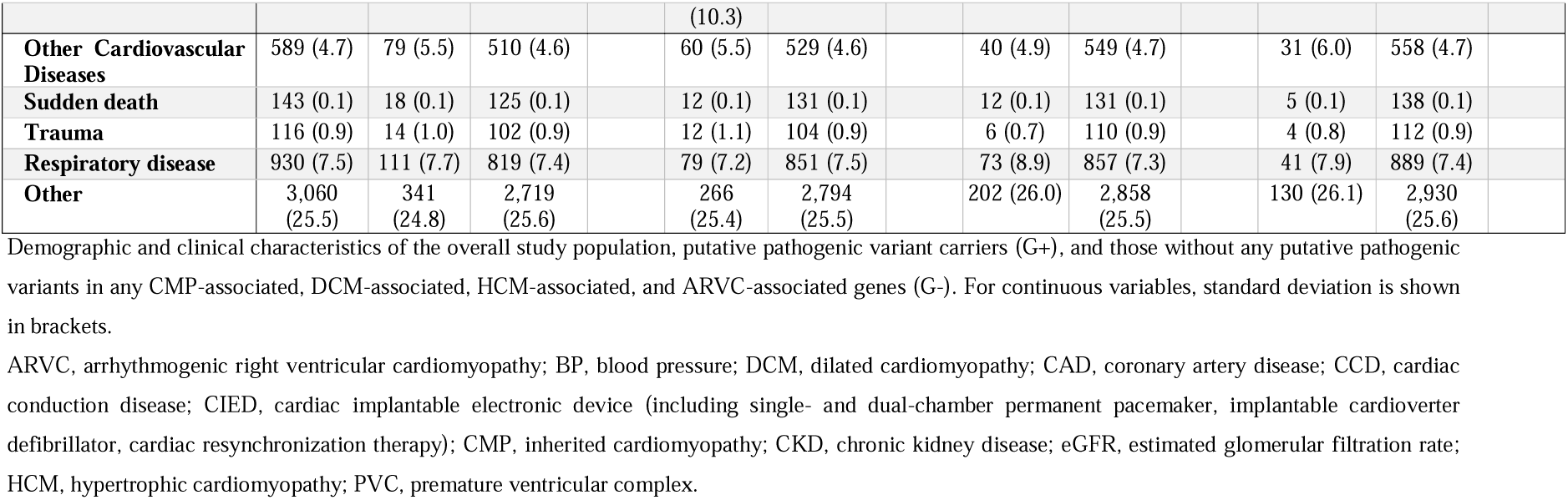
Summary of demographics and clinical characteristics.

PuPV were found in all candidate genes. The most common affected genes were, *MYH6* (n=1,732, 7.73%), *OBSCN* (n=1,712, 7.64%), *SCN5A* (n=1,498, 6.69%), *MYH7* (n=1,247, 5.57%), *TTN* (*TTNtv*, n=1,235, 5.51%), and *FLNC* (n=1,193, 5.33%). There were 358 (1.60%) individuals with PuPV in *LMNA* and 1,519 (6.8%) individuals hosted >1 PuPV in >1 CMP-associated genes (**Tables S3-6**).

### Impact of CMP-associated PuPVs on all-cause mortality

In 2,443,309 person-years of follow-up (mean 12.2 ±1.7 years), there were only 492 (0.24%) patients lost to follow-up (48 G+ and 444 G-). Time-to-event analysis, adjusted for age and sex, demonstrated significantly higher all-cause mortality in G+ compared to G- individuals, irrespective of clinical diagnoses [HR 1.07 (95%CI 1.02-1.13; p=0.011; **Figure 1**)]. Both DCM-G+ [HR 1.08 (95%CI 1.02-1.15; p=0.014)] and HCM-G+ individuals [HR 1.08 (95%CI, 1.01-1.16; p=0.03)] displayed increased mortality compared to G- subjects. For ARVC-G+, however, there was no statistically difference in mortality as compared to G- individuals [HR 1.07 (95%CI 0.98-1.16; p=0.15)].

**Figure 1.**
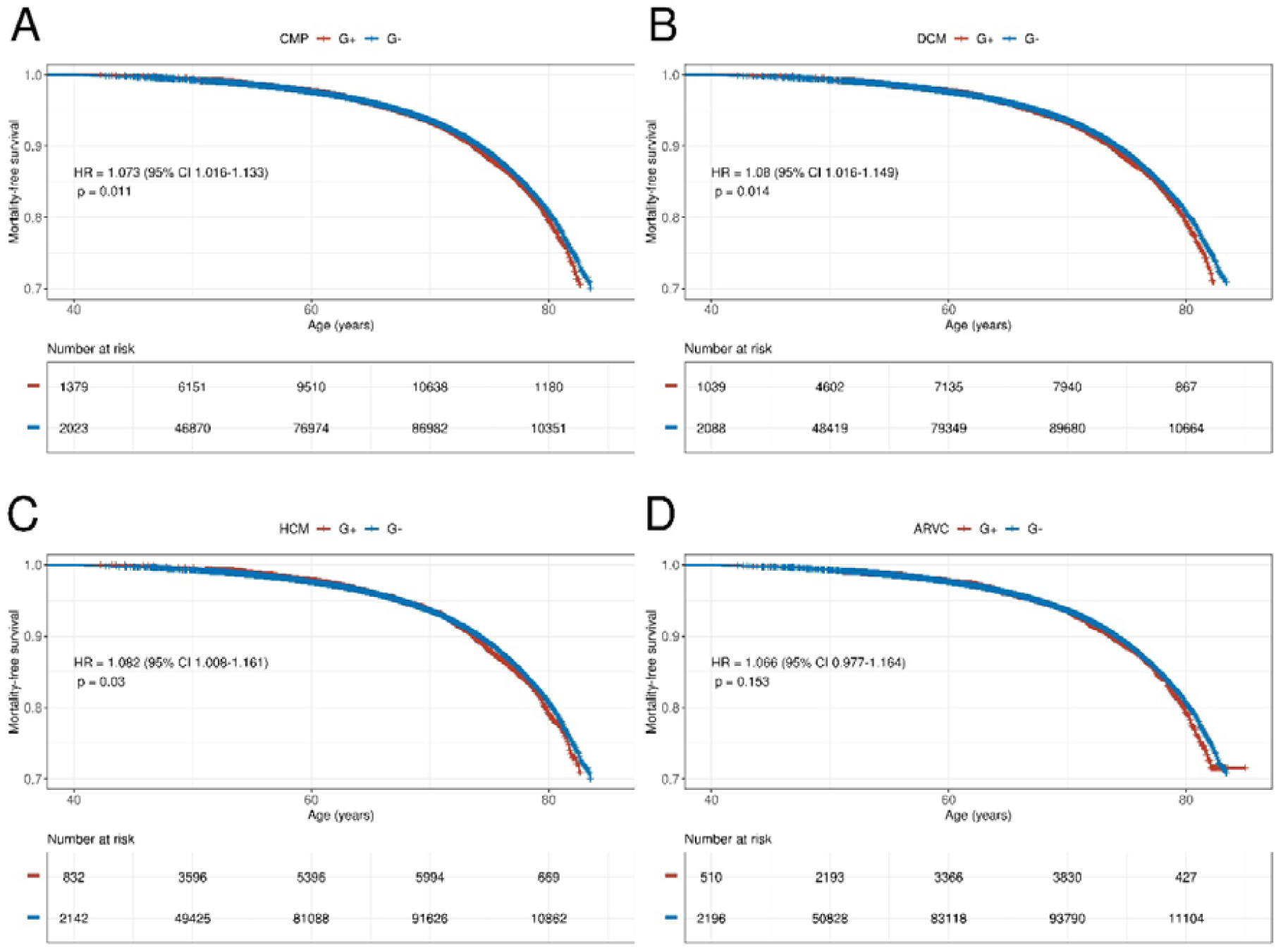
Survival analysis using Kaplan-Meier plots. Kaplan-Meier plot comparing all-cause mortality-free survival in individuals with putative pathogenic variants (G+) in any cardiomyopathy gene (A), any DCM-associated gene(s) (B), any HCM-associated gene(s) (C), and any ARVC-associated gene(s) (D) compared to those without variants in the respected group of genes (G-). Hazard ratios are displayed, calculated using Cox proportional hazard regression, corrected for sex and using age as timescale.

When splitting the CMP-associated genes by their ClinGen-asserted evidence levels (definitive/strong vs moderate vs limited evidence (**Figure S1**)), we found that the increase in mortality within the CMP-G+ individuals compared to G- controls was driven by the definitive/strong and limited groups [HR 1.09 (95%CI 1.00-1.16; p=0.03) and HR 1.09 (95%CI 1.00-1.18; p=0.04), respectively]; whereas the moderate evidence genes showed no significant impact on mortality [HR 0.83 (95% CI 0.63-1.19; p=0.23)]. This finding was similar across the individual CMPs. In fact, for HCM, individuals with PuPV in moderate group genes displayed reduced mortality compared to G- individuals [HR 0.63 (95%CI 0.41-0.95; p=0.028) (**Table S7**)]. The rates of sudden death were not significantly different between G+ and G- participants for CMP-associated genes (p=0.684); there were no significant differences when analyzed for each CMP separately either.

When limiting the analysis to the 2019 HRS high-risk ACM genes, HRS-ACM-G+ individuals did not have increased risk of mortality compared to the rest of the studied population (HR 1.08 [95%CI 0.97-1.19; p=0.029]). Participants with PuPV in ACMG actionable CMP genes (ACMG-G+) displayed increased risk of mortality compared to those with ACMG-G- status [HR 1.09 (95%CI 1.01-1.17; p=0.0255)], but there was no increase in the risk of mortality when comparing the risk in actionable ACMG G+ subjects to other CMP G+ individuals.

### Impact of CMP-associated PuPVs on developing clinical CMP

With all CMP genes grouped together, G+ status was associated with markedly increased risk of developing clinical CMP [HR 2.37 (95%CI 1.98-2.85; p<0.0001)], and when limiting to definitive/strong category genes, this rate only increased [3.24 (95%CI 2.63-4.00; p<0.0001)] (**Figures 2 and 3, Table S8**).

**Figure 2.**
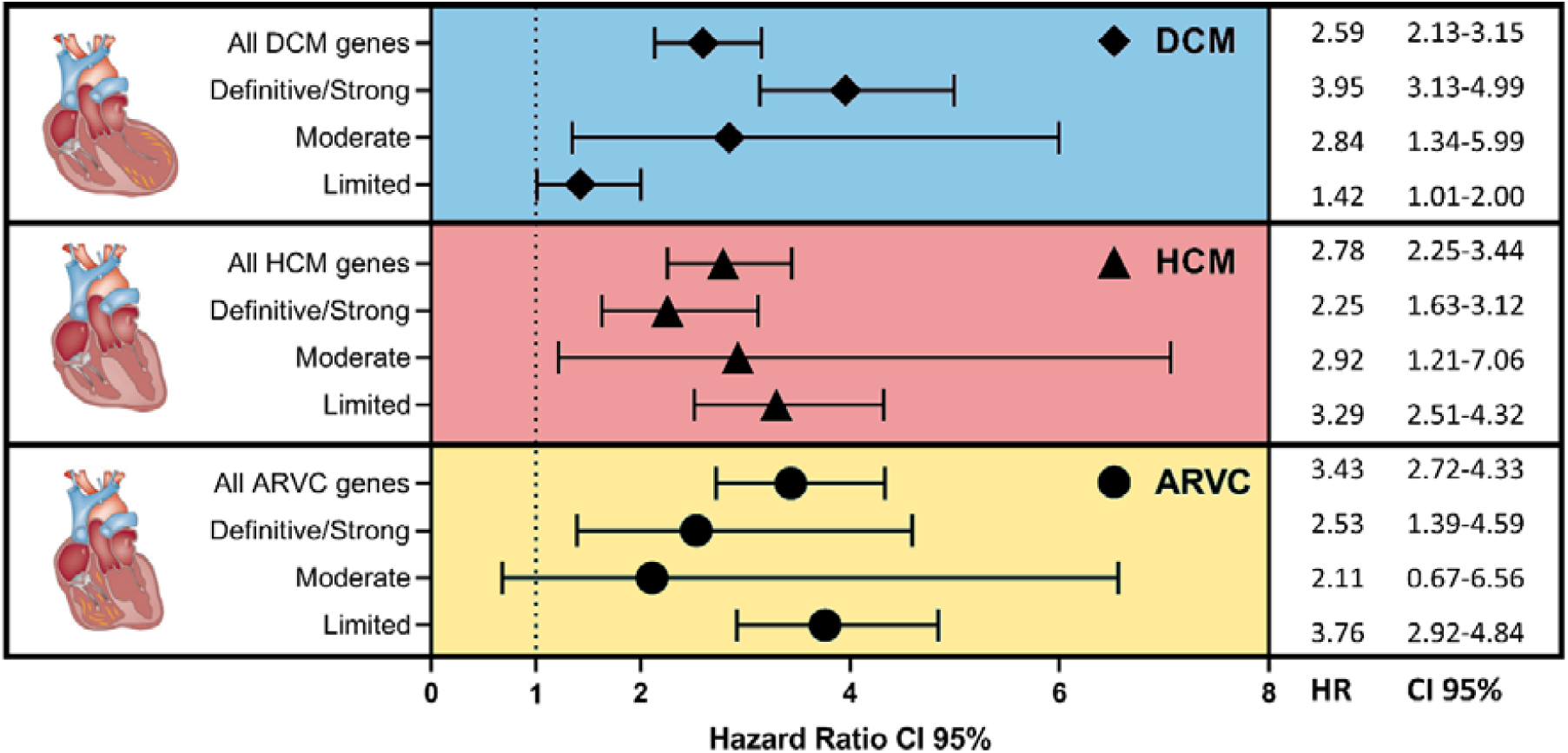
Forest plots for the development of clinical cardiomyopathy predicted by cardiomyopathy genotype. Genes implicated in cardiomyopathies are grouped by their ClinGen Gene Curation Expert Panel-asserted evidence levels. DCM, dilated cardiomyopathy; HCM, hypertrophic cardiomyopathy; ARVC, arrhythmogenic right ventricular cardiomyopathy; HR, hazard ratio; CI, Confidence interval.

**Figure 3.**
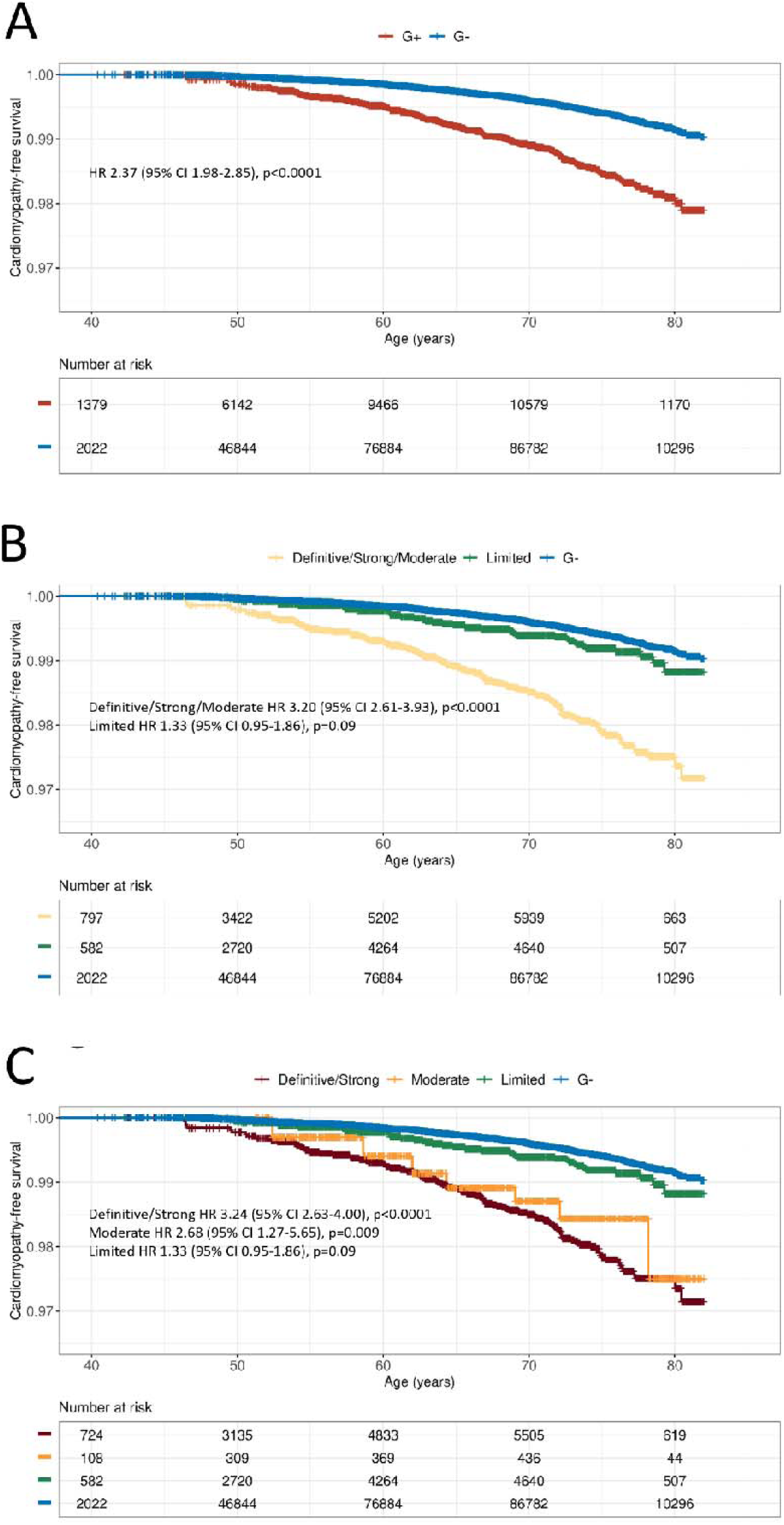
Kaplan-Meier plots showing the risk of a subsequent clinical diagnosis of cardiomyopathy. Kaplan-Meier plots comparing the risk of developing clinical cardiomyopathy in individuals with putative pathogenic variants (G+) in any cardiomyopathy gene and those without (G-) (A), and as grouped by their ClinGen-asserted evidence levels (B and C). The highest risk is in individuals with putative pathogenic variants in definitive/strong evidence genes, followed by the moderate category, then limited.

### Impact of CMP-associated PuPVs on composite clinical outcomes

The composite outcome of HF, diagnosis of CMP, stroke, CIED insertion, AF, sustained and non-sustained ventricular tachyarrhythmias was reached significantly more frequently in CMP G+ compared to G- individuals (HR 1.11 (95%CI 1.06-1.15; p<0.0001; **Figure 4**). G+ individuals for all three inherited CMP groups also displayed increased risk of developing the composite outcome [DCM-G+ HR 1.13 (95%CI 1.08-1.18; p<0.0001); HCM-G+ HR 1.13 (95%CI, 1.07-1.18; p<0.0001); and ARVC-G+ HR 1.18 (95%CI 1.11-1.25; p<0.0001)].

**Figure 4.**
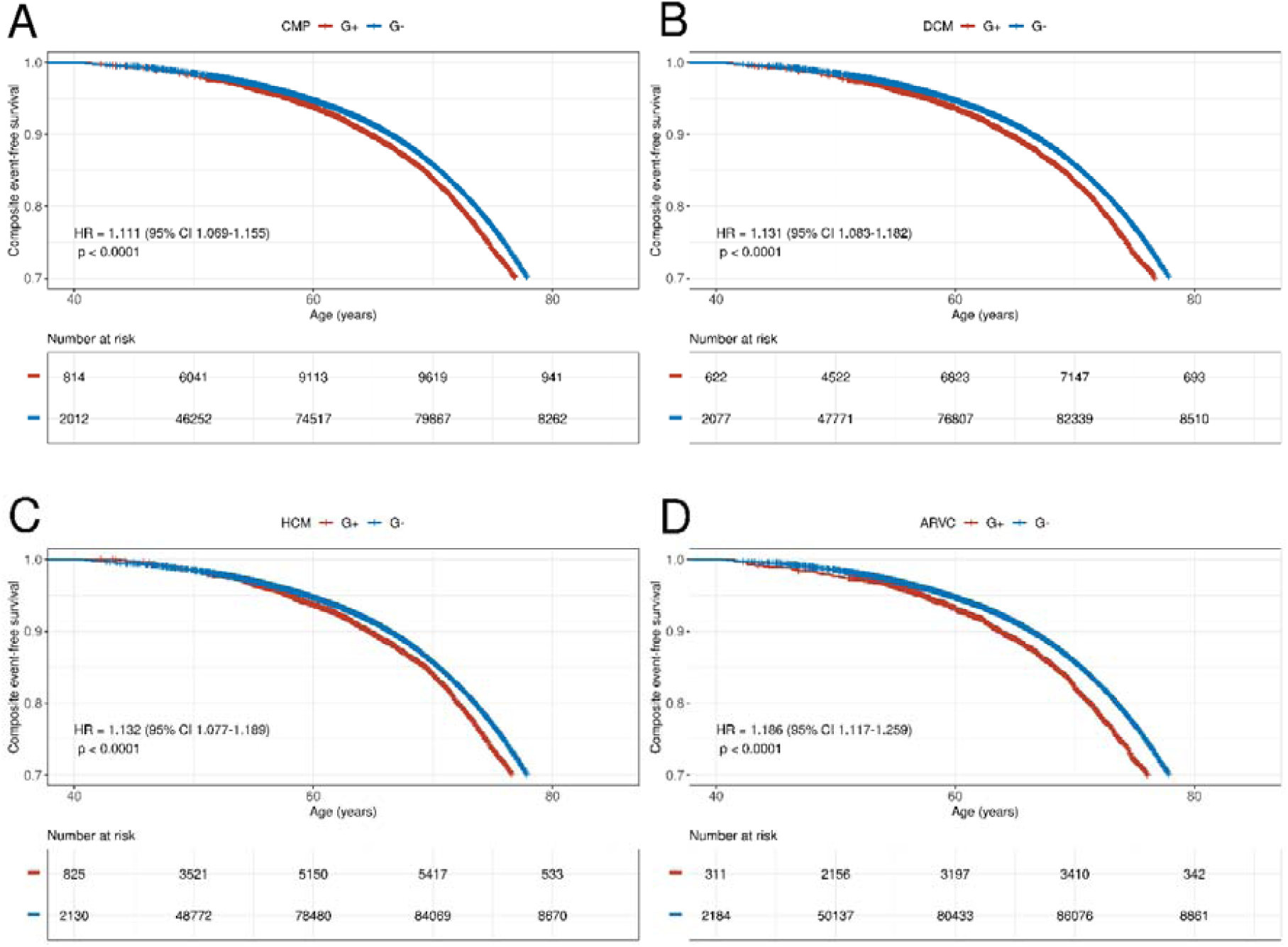
Composite outcomes analysis using Kaplan-Meier plots. Kaplan-Meier plots comparing the composite outcome-free survival (development of cardiomyopathy, heart failure, stroke, atrial fibrillation, ventricular arrhythmias, cardiac implantable electronic device insertion, death) in individuals with putative pathogenic variants (G+) in any cardiomyopathy gene (A), any DCM-associated gene(s) (B), any HCM-associated gene(s) (C), and any ARVC-associated gene(s) (D) versus those without variants in the respected group of genes (G-). Hazard ratios are displayed, calculated using Cox proportional hazard regression, corrected for sex and using age as timescale.

When analyzing the composite outcome in CMP-associated genes, PuPV in the definitive/strong evidence genes showed association with increased risk of developing the composite outcomes [1.14 (95%CI 1.08-1.20; p<0.0001)] as did PuPV in limited evidence genes [HR 1.09 (95%CI 1.03-1.15; p= 0.002)] (**Table S9**). PuPV in moderate evidence genes did not show a significant impact on composite outcomes [HR 0.97 (95%CI 0.80-1.17; p=0.78)]. A similar outcome was found for DCM-associated genes; however, for HCM and ARVC-associated genes only the limited groups had significantly increased risk of reaching the composite outcomes [HR 1.24 (95%CI 1.16-1.33; p<0.0001) and 1.23 (95%CI 1.15-1.32; p<0.0001), respectively (**Figure S2, Table S9**)]. **Figure 5** shows a heatmap with the cardiomyopathy-associated genes and the observed clinical features and outcomes.

**Figure 5.**
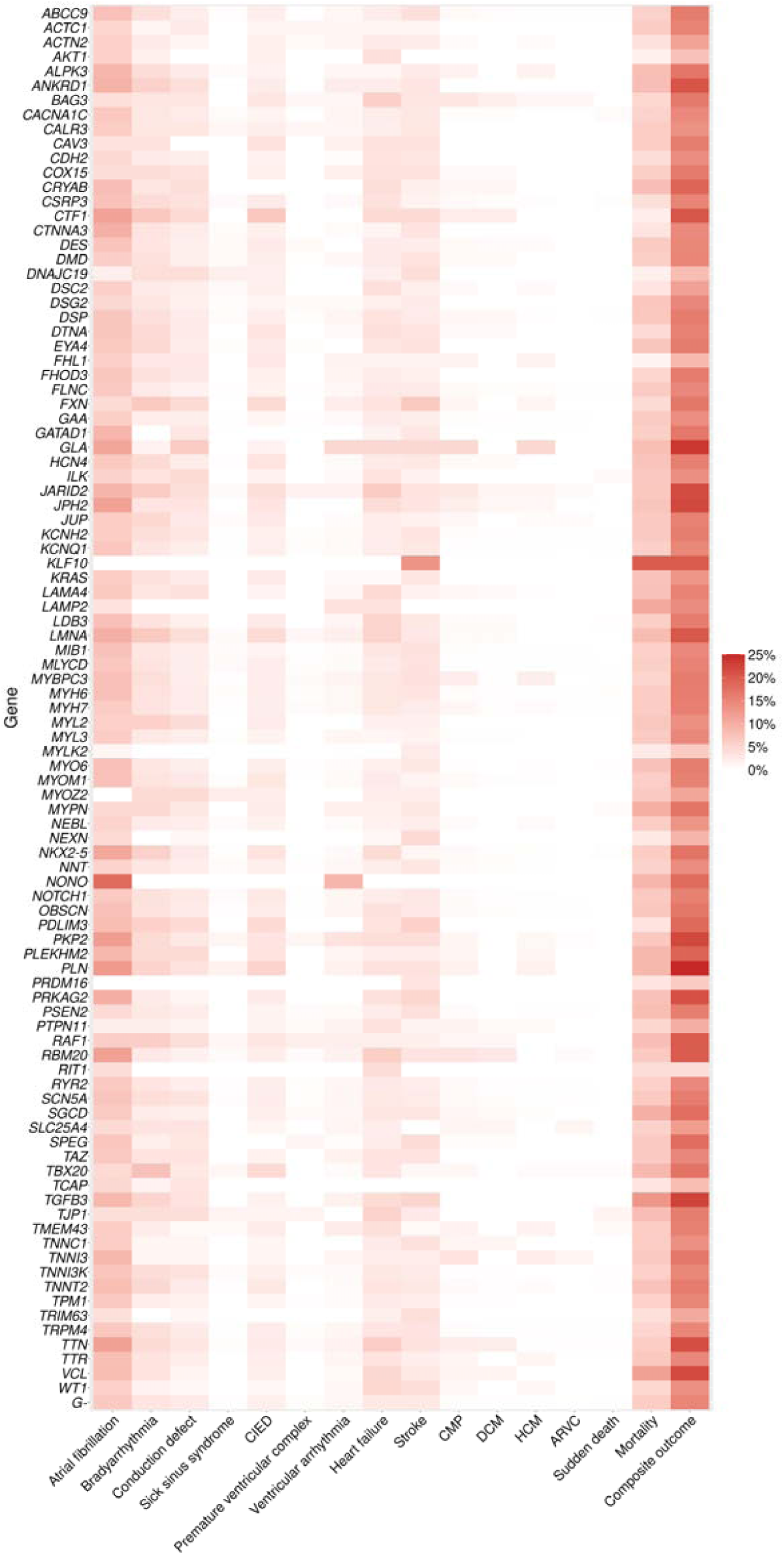
Heatmap showing the cardiomyopathy-associated genes and the clinical features and outcomes observed. ARVC, arrhythmogenic right ventricular cardiomyopathy; CIED, cardiac implantable electronic device; CMP, cardiomyopathy; DCM, dilated cardiomyopathy; HCM, hypertrophic cardiomyopathy.

When assessing the influence of the G+ state on the individual components of the composite outcome, there was increased risk of atrial fibrillation, CIED insertion, HF, and ventricular arrhythmias for nearly all groups of CMP-associated genes as classified by their ClinGen-asserted evidence levels. There was no group with increased risk of stroke (**Table S10)**. HRS-ACM G+ and ACMG G+ both had increased risk of developing the composite outcome versus G- [HR 1.10 (95%CI 1.03-1.19; p=0.004) and HR 1.15 (95%CI 1.09-1.21; p<0.0001) respectively], but not when compared with other CMP G+ (**Figure S3**). **Table 2** details the Wilson and Jungner Criteria for Genetics Population Screening and Dobrow Principles as if applied for the cardiomyopathy genes.(28)

**Table 2.**
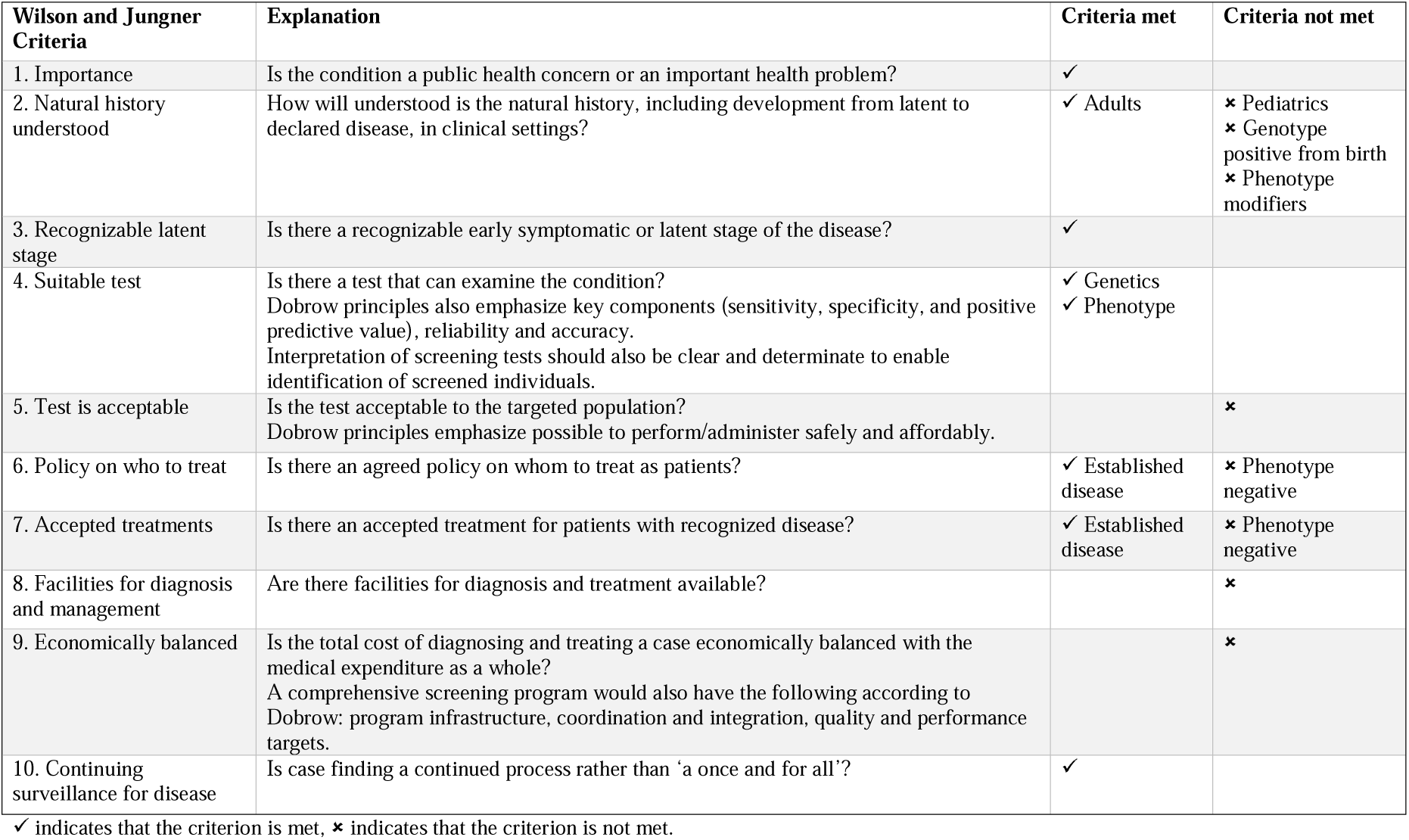
Wilson and Jungner Criteria and Dobrow principles in the context of genetics population screening of cardiomyopathy genes.

## Discussion

In an era where genetics increasingly becomes an integral part of evaluation and care of patients with genetic heart diseases,(8) the application of a ‘genotype-first’ approach to cardiomyopathies may potentially extend the clinical care from patients and families to a group of the general population at higher genetic/genomic risk of developing CMP and related outcomes. Using the UKBB prospective cohort, the largest open-access exome sequencing resource, we evaluated the impact of PuPV in CMP-associated genes on mortality, developing a CMP, and CMP-associated features in a prospective healthy cohort, and identified several important findings (**Figure 6 Central Illustration**). First, our results show that in a middle-aged White Caucasian population, CMP-G+ participants have increased all-cause mortality. Second, our study highlights a significantly increased risk of developing clinical, gene-associated CMP and other overlapping CMP phenotypes in CMP-G+ participants with three-fold increased risk in the definitive/strong evidence subgroup of genes according to ClinGen GCEP-assertions. Third, in CMP G+ individuals there is an increased risk of developing CMP-associated features/complications as compared to G- subjects. Fourth, individuals with PuPV in DCM- and/or HCM-associated genes, but not those with PuPV in ARVC-associated genes, have increased mortality as compared to those with no PuPV in the respected group of genes. Fifth, individuals with PuPV in the ACMG-defined actionable CMP-associated genes also show increased risk of mortality, but PuPV in HRS-defined high-risk ACM genes did not have an impact on mortality in the UKBB population.

**Figure 6.**
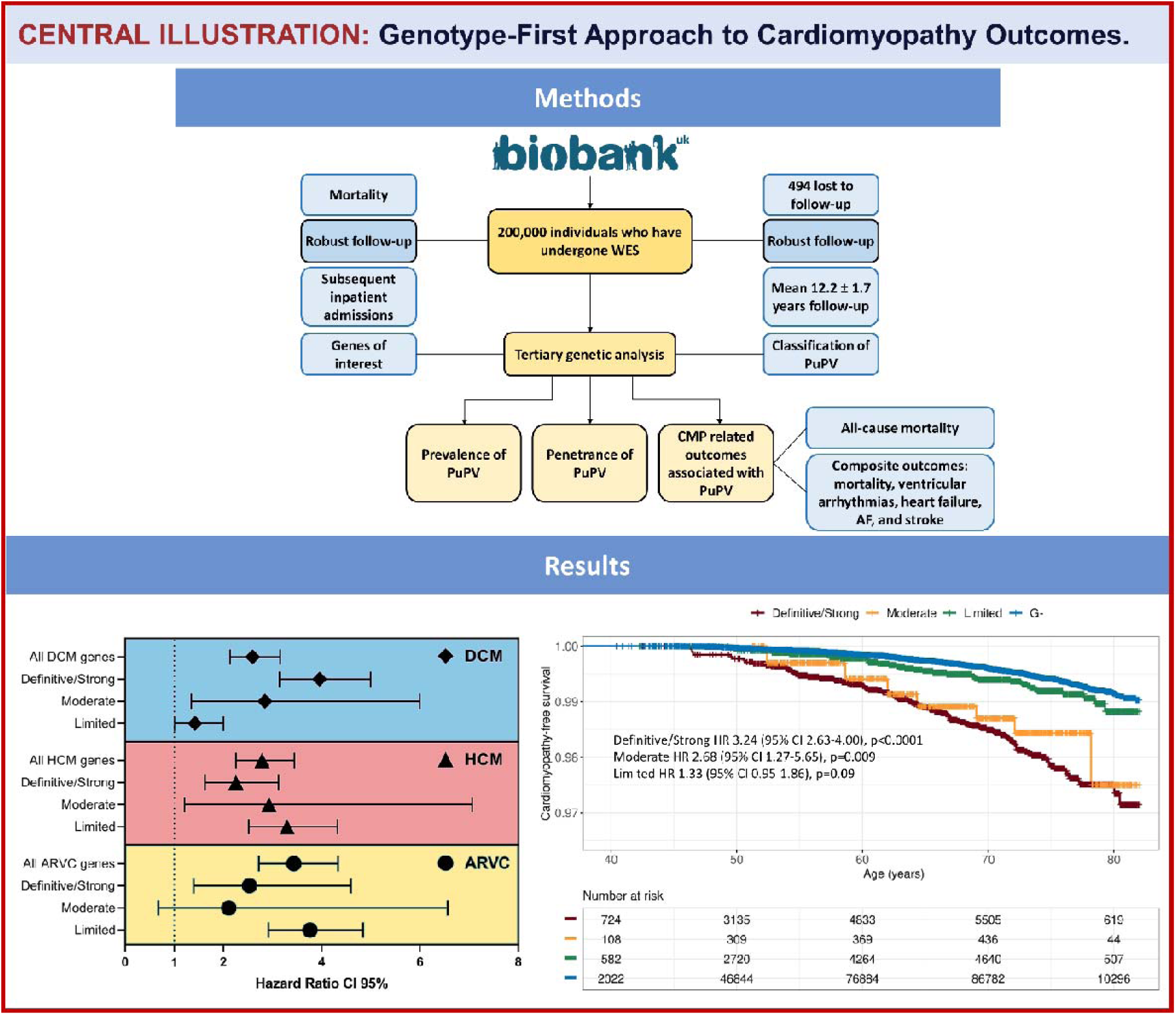
Central Illustration. CMP, cardiomyopathy; DCM, dilated cardiomyopathy; HCM, hypertrophic cardiomyopathy; ARVC, arrhythmogenic cardiomyopathy; PuPV, putative pathogenic gene variant; AF, atrial fibrillation; WES, whole exome sequencing; CI, Confidence interval.

The paradigm of cardiomyopathies is largely based on classic Mendelian disease with the majority of cases being autosomal dominant and adult onset.(2) This mode of inheritance and development of phenotype at adulthood, usually at middle age for DCM and HCM, allows these PuPV to be passed on to the next generation, i.e. most individuals have already procreated offspring, before being diagnosed with disease, which may in part explain the higher prevalence of potentially deleterious variants in individuals with no clinical features of disease. Next, the concept of CMP being only Mendelian, ‘one gene, one disease’ is being challenged with the exception of some forms, given the wide variation with penetrance and expressivity, suggesting environmental factors, as well as other genetic modifiers.(29,30) Genetic modifiers include biallelic compound heterozygotes, recessive PuPV carriers, as well as digenic inheritance. However, data also suggest polygenic contributions with SNPs.(11) Limited data also suggest that PuPV might be more frequent in CMP-associated genes than the disease prevalence itself, highlighting the critical role of knowledge to interpret incidental findings and further underscoring the role of phenotype modulators.(31) The combination of genetic effect modifiers and environmental factors may explain why some individuals can show markedly more severe phenotype than individuals in the same family carrying the same PuPV, as well as differences in disease severity compared to unrelated individuals.

The significant increase in developing clinical CMP and overlapping CMPs in CMP-G+ participants highlight the causal role of CMP gene PuPVs in these cardiomyopathies and the likelihood of developing the phenotype, while providing further data supporting low clinical penetrance of these PuPVs at population level. Our further analysis revealed a clear difference in outcome based on the classification of the genes based on the ClinGen GCEPs as individuals with PuPV in definitive/strong evidence level genes had the highest hazard ratio with narrow confidence intervals. Interestingly, individuals with PuPV in limited evidence genes had higher mortality rates, indicating that some of these genes might underlie rather severe disease phenotypes/complications with fatal course. Any given classification is representative only of the level of evidence at the time of curation, thus, evidence level is likely to change over time. It is notable that one of the genes only recently emerged as a cause of autosomal dominant HCM, *ALPK3*, was classified to limited evidence category in 2018, but a more recent publication provided a strong evidence of causality,(32) and will likely result in reclassification of this gene into the moderate or strong category in an ongoing curation of the ClinGen Hereditary Cardiovascular Disease Gene Curation Expert Panel. Thus, while the evidence regarding disease causality of PuPV in certain genes might be limited, this at times may reflect the fact that these genes are only newly discovered and not studied in large CMP cohorts, similar to *ALPK3*. This ultimately holds significance for any gene in any future endeavors for population level screening.

The increase in CMP-associated features in G+ participants is of importance in clinical settings as the paradigm of CMP is based on the extreme phenotype of HCM, DCM or ARVC. Experts who routinely care for families with CMP have reported milder and earlier phenotypic forms, in part because of more aggressive screening with multimodality imaging and arrhythmia assessment, while recognizing there are *forme frustes* of disease.(33) The fact that PuPVs in these known causal genes, with Mendelian autosomal dominant inheritance pattern, can contribute to mortality risk and composite outcomes, without showing overt structural disease meeting DCM, HCM or ARVC criteria is the major finding of our study. While the reasons behind the increased all-cause mortality in CMP-G+ individuals mostly without clear diagnosis of CMPs is unclear, one reason responsible for higher mortality can be malignant ventricular arrhythmias prior to the development of detectable CMP phenotype.(34) Studies found pathogenic and likely pathogenic variants in CMP genes in patients with unexplained cardiac arrest, highlighting the arrhythmogenic potential of such variants in the absence of apparent CMP features.(35,36) Other effect modifiers in the clinical setting include exercise and inflammation, which might promote development of phenotype or CMP-related complications.(30) However, the increased mortality may not necessarily be due to higher rates of sudden cardiac death—it is all-cause in this study, and this warrants further investigation.

Our study also shows that ARVC G+ individuals show no difference in all-cause mortality. One potential explanation might be that while DCM and HCM in relevant proportion of cases present between fifth and seventh decades of life (the age of this cohort), ARVC typically presents over 10-20 years earlier.(3) These highly penetrant cases may present with sudden cardiac death as a sentinel event, reflecting survival bias in the UKBB participants. The UKBB also has healthy volunteer bias, as shown by CMR-based studies thus far.(37) When using the 2019 HRS grouping of high risk ACM genes, this also did not show increased mortality in this group, which is contrary to some of these genes being reported as penetrant and potentially lethal in *patient cohorts*. This might reflect survival bias or small sample size (as compared to DCM-G+ and HCM-G+ groups the ARVC-G+ cohort was much smaller). The ACMG actionable CMP-related genes are a smaller number of genes, selected based on high penetrance, and potentially lethal course. In the UKBB cohort, PuPV in these genes were associated with higher all-cause mortality, supporting the ACMG recommendations for actionability.(15) It should be noted the ACMG recommendations for actionability are based on incidental findings in these genes and not specifically to population-based screening, because of a lack of sufficient evidence.

Our findings of increased mortality and higher rate of developing CMP-associated features in most CMP G+ groups in the general population provide further data in the field of ‘genotype-first’ approach, and support the studies by de Marvao *et al*, who recently reported low penetrance of HCM yet increased risk of developing HCM-related features in individuals with sarcomeric PuPV.(33) We also confirm that the presence of PuPV in CMP-associated genes is associated with higher rate of atrial fibrillation/flutter, confirming the findings of Yoneda *et al*(38) from the TOPMed cohort (145 CMP genes) and Haggerty *et al*. from the Geisinger MyCode Community Health Initiative and the PennMedicine BioBank (*TTVtv*).(39) Furthermore, we demonstrate higher mortality, higher prevalence of ventricular arrhythmias, CIED insertions, and HF in all CMP-G+ groups, in line with the data from clinical cardiomyopathy cohorts.(40) Overall, these data suggest that CMP-PuPV might result in reduced ‘cardiovascular reserve’, whereby genetically affected individuals are susceptible to development of CMP-related adverse outcomes with the influence of otherwise mild effect modifier(s), such as myocarditis,(30,41) endurance exercise,(42) alcohol consumption,(43) and other potentially under-recognized/under-investigated effect modifiers (e.g. race and ethnicity).

Wilson and Junger described 10 principles that screening programs should meet to be successful, which can be applied to a population genetic screening program for PuPV in CMP-associated genes.(44) Dobrow et al.(45) consolidated principles for screening, and both Wilson-Jungner criteria and Dobrow principles are summarized in **Table 2**. Several of these criteria are met: criteria (1) importance, (3) a recognizable latent stage; (4) a suitable test; and (10) need for continuing surveillance. Several criteria are partially met: criterion (2) regarding natural history of CMPs is understood for adult-onset disease but not as much in pediatric forms of CMP; criterion (6) regarding policy and criterion (7) on acceptable treatments are partially met for established disease, such as cardiomyopathies, heart failure and arrhythmias but remain unclear for genotype-positive/-phenotype-negative individuals. Criterion (8) regarding the availability of facilities for diagnosis and (9) for the screening methodology being economically balanced, are not met yet. This applies to clinical evaluation for phenotyping, such as the use of multimodality imaging, necessary for early detection and accurate phenotyping, especially given pathogenic variants in certain genes can cause multiple and overlapping CMP phenotypes.(46) At present, the cost of genetic screening may outweigh the benefits provided, although formal cost-effectiveness analyses specific to cardiovascular disease are needed. The falling costs of genetic sequencing (both WES/WGS), coupled with robust analytical pipelines, may make this achievable in the future. The goal of a $100 genome in the next decade may make this a feasible solution, though this should be tempered with adequate safeguards and robust evidence. There are serious risks of ‘genetic discrimination’ which may be based purely speculatively on incidental variants of uncertain significance.(47) Lastly, case finding should be a continuous process, which would likely involve genetic screening of a cohort once they reach a certain age.

## Strengths and Limitations

Strengths of our study include (a) large sample size; (b) access to a general population; (c) small number of participants lost to follow-up/withdrawal; (d) long duration of follow-up; and (e) robust and conservative variant annotation pipeline. One limitation of our study is that the UKBB only includes individuals aged 40-69 years old at enrollment. This means it is likely that patients with highly penetrant variants, with younger age at presentation, and high risk of sudden cardiac death at young ages and as sentinel manifestation, are not included in this cohort. Secondly, the UKBB is affected by healthy volunteer bias.(48) Thirdly, the UKBB mainly has white British participants, which limits generalizability of these findings; especially as it is known that the genetic architecture of CMPs varies by ethnicity.(49,50). Fourth, we used ClinGen GCEP-asserted gene panels to define the gene list. Since the evidence regarding gene-disease causality extends very quickly, it is possible some of the genes included may be classified into a different evidence category at re-curation.

## Conclusions

In an adult general population, the presence of PuPV in CMP-associated genes predicts all-cause mortality, subsequent risk of developing cardiomyopathy and composite CMP-related outcomes, irrespective of ultimate phenotype. This suggests a potential role for genetic screening of PuPV variants in CMP-associated genes to inform cardiovascular outcomes and mortality and to transform personalized medicine to population-level precision health. While the mechanisms behind increased mortality in individuals with PuPV in cardiomyopathy genes remains unclear and requires further investigations, PuPV might result in reduced ‘cardiovascular reserve’, whereby other effect modifiers promote adverse outcomes at a lower threshold. Further work on identification of variants with larger effect size, cost-effectiveness analyses and understanding how early measures can ameliorate the genetic risk are required before general population screening for CMP-associated PuPVs can be recommended.

## Perspectives

### Competency in Medical Knowledge

Inherited cardiomyopathies show variable and heterogeneous phenotypes, with severe cases manifesting first in first three decades of life. Performing genetic testing on patients and their relatives is a valuable tool to inform diagnosis, prognostication and management strategy.

### Translational Outlook 1

Using a ‘genotype first’ approach for population-level screening in cardiomyopathy-associated genes can help identify those at risk for developing cardiomyopathy features, or related complications regardless of clinical phenotype status.

### Translational Outlook 2

Before population level screening can be implemented, additional advances in genetic sequencing technologies are crucial to further decrease the cost of genetic testing, and more research is needed to identify and characterize genes and variants with large effect size, and to develop strategies to ameliorate the genetic risk of developing cardiomyopathy or related outcomes.

## Data Availability

All data produced in the present study are available upon accepted application to access the UK Biobank data.

## Acknowledgement

We thank all the UK Biobank participants.

## Abbreviations

AF: Atrial fibrillation or flutter
ARVC: Arrhythmogenic right ventricular cardiomyopathy
CCD: Cardiac conduction disease
CIED: Cardiac implantable electronic device
ClinGen: Clinical Genome Resource
CMP: Cardiomyopathy
DCM: Dilated cardiomyopathy
FAF: Filtering allele frequency
GCEP: Gene Curation Expert Panel
HCM: Hypertrophic cardiomyopathy
HF: Heart Failure
pLOF: predicted loss-of-function
PuPV: Putative pathogenic gene variant
UKBB: UK Biobank
WES: Whole exome sequencing

