## Supplemental Material for "Screening for Pathogenic Variants in Cardiomyopathy Genes Predicts Mortality and Composite Outcomes in UK Biobank"

Short title: Genotype-First Approach to Cardiomyopathy Outcomes

Babken Asatryan, MD, PhD; Ravi A. Shah, MB BChir; Ghaith Sharaf Dabbagh, MD;

Andrew P. Landstrom, MD, PhD; Dawood Darbar, MD, MBChB; Mohammed Y Khanji, MBBCh, PhD; Luis R. Lopes, MD, PhD; Stefan van Duijvenboden, PhD; Daniele Muser, MD;

Aaron Mark Lee, BSc(hons), MBBS, PhD; Christopher M. Haggerty, PhD; Pankaj Arora, MD; Christopher Semsarian, MBBS, PhD, MPH; Tobias Reichlin, MD; Virend K. Somers, MD, PhD;

Anjali T. Owens, MD; Steffen E. Petersen, MSc, MPH, MD, DPhil; Rajat Deo, MD, MPH;

Patricia B Munroe, MSc, PhD; Nay Aung, MBBS, PhD, MRCP; C. Anwar A. Chahal, MBChB, PhD, MRCP, on behalf of the Genotype-First Approach Investigators

**Supplemental Material**

**Supplemental methods**

*Selection of genes of interest*

The ClinGen DCM panel includes 11 genes with definitive evidence (*BAG3, DES, FLNC, LMNA, MYH7, PLN, RBM20, SCN5A, TNNC1, TNNT2, TTN*), one with strong evidence (*DSP*), seven with moderate evidence (*ACTC1, ACTN2, JPH2, NEXN, TNNI3, TPM1, VCL*) and 25 with limited evidence for causality (*ABCC9, ANKRD1, CSRP3, CTF1, DSG2, DTNA, EYA4, GATAD1, ILK, LAMA4, LDB3, MYBPC3, MYH6, MYL2, MYPN, NEBL, NKX2-5, OBSCN, PLEKHM2, PRDM16, PSEN2, SGCD, TBX20, TCAP, TNNI3K*) (1). The HCM panel includes 8 definitive evidence genes (*MYBPC3, MYH7, TNNT2, TNNI3, TPM1, ACTC1, MYL3, MYL2*), 3 moderate evidence genes (*CSRP3, TNNC1, JPH2*), and 16 low evidence genes (*TTN, KLF10, MYPN, ANKRD1, MYLK2, MYOZ2, NEXN, VCL, TRIM63, RYR2, MYH6, OBSCN, PDLIM3, TCAP, MYOM1, CALR3*). In addition, the syndromic genes were also included, with 20 definitive evidence genes (*PLN, CACNA1C, DES, FHL1, FLNC, GLA, LAMP2, PRKAG2, PTPN11, RAF1, RIT1, TTR, ABCC9, BAG3, CAV3, CRYAB, FXN, GAA, MYO6, SLC25A4*), 2 strong evidence genes (*ALPK3, COX15*), and 2 moderate evidence genes (*ACTN2, LDB3*) (2). The ARVC panel includes 6 strong evidence genes (*PKP2, DSP, DSG2, DSC2, JUP, TMEM43*), 2 moderate evidence genes (*PLN, DES*), and 10 limited evidence genes (*SCN5A, LMNA, CDH2, CTNNA3, TGFB3, TTN, TJP1, MYH7, MYBPC3, MYL3*).

Genes included in the Heart Rhythm Society expert consensus statement on arrhythmogenic cardiomyopathies were (3,4):

The American College of Genetics and Genomics latest actionable list includes 78 genes, of which X are cardiomyopathy genes. These were also assessed as a separate group and included: *MYBPC3, MYH7, TNNT2, TNNI3, TPM1, MYL3, ACTC1, PRKAG2, GLA, MYL2, LMNA, FLNC, TTN, PKP2, DSP, DSC2, TMEM43, DSG2, GAA, RYR2, KCNQ1, KCNH2, SCN5A*, *TNNC1*, *RBM20*, *BAG3*, *DES*, and *TTR*.

**Table S1. Table of phenotypic definitions used.**

| **Phenotype** | **Data fields** | **Field names** | **Data codes** | **Data code definitions** |
| --- | --- | --- | --- | --- |
| Arrhythmogenic right ventricular cardiomyopathy | 41202  41204  40001  40002 | Diagnoses – main ICD10  Diagnoses – secondary ICD10  Underlying (primary) cause of death: ICD10  Contributory (secondary) cause of death: ICD10 | I42.8 | Other cardiomyopathies |
| Atrial fibrillation or flutter | 20002 | Non-cancer illness code, self-reported | 1471, 1483 | Atrial fibrillation, Atrial flutter |
| Atrial fibrillation or flutter | 20004 | Operation code, self-reported | 1524 | Cardioversion |
| Atrial fibrillation or flutter | 41202  41204  40001  40002 | Diagnoses – main ICD10  Diagnoses – secondary ICD10  Underlying (primary) cause of death: ICD10  Contributory (secondary) cause of death: ICD10 | I48, I48.0, I48.1, I48.2, I48.3, I48.4, I48.9 | Atrial fibrillation and flutter, Paroxysmal atrial fibrillation, Persistent atrial fibrillation, Chronic atrial fibrillation, Typical atrial flutter, Atypical atrial flutter, Atrial fibrillation and atrial flutter, unspecified |
| Atrial fibrillation or flutter | 41203  41205 | Diagnoses – main ICD9  Diagnoses – secondary ICD9 | 4273 | Atrial fibrillation and flutter |
| Atrial fibrillation or flutter | 41200  41210 | Operative procedures – main OPCS Operative procedures – secondary OPCS | K57.1, K62.1, K62.2, K62.3, K62.4 | Percutaneous transluminal ablation of atrioventricular node, Percutaneous transluminal ablation of pulmonary vein to left atrium conducting system, Percutaneous transluminal ablation of atrial wall for atrial flutter,  Percutaneous transluminal ablation of conducting system of heart for atrial flutter NEC, Percutaneous transluminal internal cardioversion NEC |
| Bradyarrhythmias | 20002 | Non-cancer illness code, self-reported | 1486 | Sick sinus syndrome |
| Bradyarrhythmias | 41202  41204  40001  40002 | Diagnoses – main ICD10  Diagnoses – secondary ICD10  Underlying (primary) cause of death: ICD10  Contributory (secondary) cause of death: ICD10 | I44, I44.1, I44.2, I44.3, I44.5, I49.5 | Atrioventricular and left bundle-branch block, Atrioventricular block, second degree, Atrioventricular block, complete, Other and unspecified atrioventricular block, Other specified heart block, Sick sinus syndrome |
| Bradyarrhythmias | 41203  41205 | Diagnoses – main ICD9  Diagnoses – secondary ICD9 | 4260, 4261, 4266 | Atrioventricular block, complete, Atrioventricular block, other and unspecified, Other specified heart block |
| Bradyarrhythmias | 20004 | Operation code, self-reported | 1096, 1548, 1549 | Pacemaker/defibrillator insertion, Pacemaker insertion, Pacemaker battery change |
| Bradyarrhythmias | 3079 | Pace-maker, verbal interview | 1 | Yes |
| Bradyarrhythmias | 41200  41210 | Operative procedures – main OPCS Operative procedures – secondary OPCS | K60, K60.1, K60.2, K60.3, K60.4, K60.5, K60.6, K60.8, K60.9, K61, K61.1, K61.2,  K61.3, K61.4, K61.5, K61.6, K61.8, K61.9 | Cardiac pacemaker system introduced through vein, Implantation of intravenous cardiac pacemaker system NEC, Resiting of lead of intravenous cardiac pacemaker system, Renewal of intravenous cardiac pacemaker system, Removal of intravenous cardiac pacemaker system, Implantation of intravenous single chamber cardiac pacemaker system, Implantation of intravenous dual chamber cardiac pacemaker system, Other specified cardiac pacemaker system introduced through vein, Unspecified cardiac pacemaker system introduced through vein, Other cardiac pacemaker system, Implantation of cardiac pacemaker system NEC, Resiting of lead of cardiac pacemaker system NEC, Renewal of cardiac pacemaker system  NEC, Removal of cardiac pacemaker system NEC, Implantation of single chamber cardiac pacemaker system, Implantation of dual chamber cardiac  pacemaker system, Other specified other cardiac pacemaker system, Unspecified other cardiac pacemaker system |
| Conduction system diseases | 41202  41204  40001  40002 | Diagnoses – main ICD10  Diagnoses – secondary ICD10  Underlying (primary) cause of death: ICD10  Contributory (secondary) cause of death: ICD10 | I44.0, I44.4, I44.5, I44.6, I44.7, I45.0, I45.1, I45.2, I45.3, I45.4 | Atrioventricular block, first degree, Left anterior fascicular block, Left posterior fascicular block, Other and unspecified fascicular block, Left bundle-branch block, unspecified, Right fascicular block, Other and unspecified right bundle- branch block, Bifascicular block, Trifascicular block, Nonspecific intraventricular block |
| Conduction system diseases | 41203  41205 | Diagnoses – main ICD9  Diagnoses – secondary ICD9 | 4263, 4264, 4265 | Other left bundle branch block, Right bundle branch block, Bundle branch block, unspecified |
| Coronary artery disease | 41202  41204  40001  40002 | Diagnoses – main ICD10  Diagnoses – secondary ICD10  Underlying (primary) cause of death: ICD10  Contributory (secondary) cause of death: ICD10 | I21.X, I22.X, I23.X, I24.1, I25.2 | Myocardial infarction |
| Coronary artery disease | 20002 | Non-cancer illness code, self-reported | 1075 | Heart attack/myocardial infarction |
| Coronary artery disease | 41200  41210 | Operative procedures – main OPCS Operative procedures – secondary OPCS | K40.1–40.4, K41.1–41.4, K45.1–45.5, K49.1–49.2, K49.8–49.9, K50.2, K75.1–75.4, K75.8–75.9 | Coronary artery bypass grafting, coronary angioplasty with or without stenting |
| Coronary artery disease | 20004 | Operation code, self-reported | 1070, 1095 | coronary angioplasty (ptca) +/- stent, coronary artery bypass grafts (cabg) |
| Diabetes | 20002 | Non-cancer illness code, self-reported | 1220  1222  1223 | Diabetes  Type 1 diabetes  Type 2 diabetes |
| Diabetes | 41202  41204  40001  40002 | Diagnoses – main ICD10  Diagnoses – secondary ICD10  Underlying (primary) cause of death: ICD10  Contributory (secondary) cause of death: ICD10 | E10-14 | Diabetes mellitus |
| Dilated cardiomyopathy | 41202  41204  40001  40002 | Diagnoses – main ICD10 Diagnoses – secondary ICD10  Underlying (primary) cause of death:ICD10  Contributory (secondary) cause of death: ICD10 | I42.0 | Dilated cardiomyopathy |
| Heart failure | 20002 | Non-cancer illness code, self-reported | 1076 | Heart failure |
| Heart failure | 41202  41204  40001  40002 | Diagnoses – main ICD10  Diagnoses – secondary ICD10  Underlying (primary) cause of death: ICD10  Contributory (secondary) cause of death: ICD10 | I11.0, I13.0, I13.2, I25.5, I50.0, I50.1, I50.9 | hypertensive heart disease, heart failure |
| Hypertension | 20002 | Non-cancer illness code, self-reported | 1065, 1072 | Essential hypertension, hypertension |
| Hypertension | 41202  41204  40001  40002 | Diagnoses – main ICD10  Diagnoses – secondary ICD10  Underlying (primary) cause of death: ICD10  Contributory (secondary) cause of death: ICD10 | I10-5 | Hypertensive diseases |
| Hypertrophic cardiomyopathy | 41202  41204  40001  40002 | Diagnoses – main ICD10  Diagnoses – secondary ICD10  Underlying (primary) cause of death: ICD10  Contributory (secondary) cause of death: ICD10 | I42.1, I42.2 | Obstructive hypertrophic cardiomyopathy, other hypertrophic cardiomyopathy |
| Premature ventricular complex | 41202  41204  40001  40002 | Diagnoses - main ICD10  Diagnoses – secondary ICD10  Underlying (primary) cause of death: ICD10  Contributory (secondary) cause of death: ICD10 | I49.3 | Premature ventricular depolarisation |
| Supraventricular arrhythmias | 20002 | Non-cancer illness code, self-reported | 1484 | Wolff-Parkinson Wwhite / WPW syndrome |
| Supraventricular arrhythmias | 41202  41204  40001  40002 | Diagnoses - main ICD10  Diagnoses – secondary ICD10  Underlying (primary) cause of death: ICD10  Contributory (secondary) cause of death: ICD10 | I45.6 | Preexcitation syndrome |
| Supraventricular arrhythmias | 41203  41205 | Diagnoses - main ICD9  Diagnoses - secondary ICD9 | 4267 | Anomalous atrioventricular excitation |
| Supraventricular arrhythmias | 41200  41210 | Operative procedures - main OPCS  Operative procedures – secondary OPCS | K52.4,  K57.4 | Open division of accessory pathway within heart, Percutaneous transluminal ablation of accessory pathway, |
| Valvular heart disease | 41202  41204  40001  40002 | Diagnoses - main ICD10  Diagnoses – secondary ICD10  Underlying (primary) cause of death: ICD10  Contributory (secondary) cause of death: ICD10 | I34-37, I39.0-39.4, I05-08, I09.1, I09.8 | Aortic, mitral, pulmonary and tricuspid valve disorders |
| Valvular heart disease | 20002 | Non-cancer illness code, self-reported | 1078, 1488, 1584, 1585, 1489, 1586, 1587, 1490 | heart valve problem/heart murmur, mitral valve prolapse, mitral valve disease, mitral regurgitation/incompetence, mitral stenosis, aortic valve disease, aortic regurgitation/incompetence, aortic stenosis |
| Ventricular arrhythmias | 41202  41204  40001  40002 | Diagnoses - main ICD10  Diagnoses – secondary ICD10  Underlying (primary) cause of death: ICD10  Contributory (secondary) cause of death: ICD10 | I47.0, I47.2, I49.0, I46.0,  I46.1, I46.9 | Reentry ventricular arrhythmia, Ventricular tachycardia, Ventricular fibrillation and flutter, Cardiac arrest with successful resuscitation, Sudden cardiac death, so described, Cardiac arrest, unspecified |
| Ventricular arrhythmias | 41203  41205 | Diagnoses - main ICD9  Diagnoses - secondary ICD9 | 4271, 4274, 4275 | Paroxysmal ventricular tachycardia, Ventricular fibrillation and flutter, Cardiac arrest |
| Ventricular arrhythmias | 41200  41210 | Operative procedures - main OPCS  Operative procedures – secondary OPCS | K57.6, K64.1, X50.3, X50.4, X50.8, X50.9 | Percutaneous transluminal ablation of ventricular wall NEC, Percutaneous radiofrequency ablation of epicardium, Advanced cardiac pulmonary resuscitation, External ventricular defibrillation, Other specified external resuscitation, Unspecified external resuscitation |

UK Biobank data field numbers and names are referenced along with the data codes and their associated definitions where appropriate.

Table adapted from eTable 2 and 3 in Khurshid et al. 2018, additional definitions from Khera et al. 2018 and Aragam et al. 2018 where available.(5-7)

**Table S2. Summary of additional demographics and clinical characteristics.**

|  | Overall | CMP G+ | CMP G- | P | DCM G+ | DCM G- | p | HCM G+ | HCM G- | p | ARVC G+ | ARVC G- | p |
| --- | --- | --- | --- | --- | --- | --- | --- | --- | --- | --- | --- | --- | --- |
| N | 200,619 | 22,401 | 178,218 |  | 16,798 | 183,821 |  | 12,745 | 187,874 |  | 8,028 | 192,591 |  |
| Ethnicity (%) |  |  |  | <0.001 |  |  | <0.001 |  |  | <0.001 |  |  | <0.001 |
| White | 188,241 (93.9) | 20,033 (89.6) | 168,208 (94.5) |  | 15,042 (89.7) | 173,199 (94.3) |  | 11,244 (88.4) | 176,997 (94.3) |  | 7,247 (90.4) | 180,994 (94.1) |  |
| Asian or Asian  British | 4,265 (2.1) | 971 (4.3) | 3,294 (1.9) |  | 721 (4.3) | 3,544 (1.9) |  | 598 (4.7) | 3,667 (2.0) |  | 321 (4.0) | 3,944 (2.1) |  |
| Black, Black  British, Caribbean or  African | 3,230 (1.6) | 539 (2.4) | 2691 (1.5) |  | 403 (2.4) | 2,827 (1.5) |  | 351 (2.8) | 2,879 (1.5) |  | 175 (2.2) | 3,055 (1.6) |  |
| Other ethnic group | 1,929 (1.0) | 373 (1.7) | 1,556 (0.9) |  | 267 (1.6) | 1,662 (0.9) |  | 245 (1.9) | 1,684 (0.9) |  | 110 (1.4) | 1,819 (0.9) |  |
| Mixed or multiple  ethnic groups | 1,303 (0.7) | 190 (0.8) | 1,113 (0.6) |  | 139 (0.8) | 1,164 (0.6) |  | 116 (0.9) | 1,187 (0.6) |  | 71 (0.9) | 1,232 (0.6) |  |
| Not specified | 794 (0.4) | 107 (0.5) | 687 (0.4) |  | 85 (0.5) | 709 (0.4) |  | 62 (0.5) | 732 (0.4) |  | 45 (0.6) | 749 (0.4) |  |
| Chinese | 642 (0.3) | 152 (0.7) | 490 (0.3) |  | 117 (0.7) | 525 (0.3) |  | 103 (0.8) | 539 (0.3) |  | 47 (0.6) | 595 (0.3) |  |
| Systolic blood pressure (mmHg) | 139.61 (19.59) | 139.12 (19.55) | 139.68 (19.60) | <0.001 | 138.96 (19.52) | 139.67 (19.60) | <0.001 | 138.86 (19.43) | 139.66 (19.60) | <0.001 | 138.77 (19.51) | 139.65 (19.59) | <0.001 |
| Diastolic blood pressure (mmHg) | 82.17 (10.68) | 81.99 (10.73) | 82.20 (10.68) | 0.008 | 81.96 (10.74) | 82.19 (10.68) | 0.008 | 81.99 (10.76) | 82.18 (10.68) | 0.054 | 81.93 (10.77) | 82.18 (10.68) | 0.047 |
| Body mass index (kg/m^2^) | 27.38 (4.76) | 27.35 (4.75) | 27.38 (4.76) | 0.406 | 27.33 (4.72) | 27.38 (4.76) | 0.211 | 27.35 (4.79) | 27.38 (4.75) | 0.524 | 27.35 (4.70) | 27.38 (4.76) | 0.629 |
| eGFR (ml/min) | 86.99 (16.74) | 87.40 (16.84) | 86.94 (16.72) | <0.001 | 87.40 (16.91) | 86.96 (16.72) | 0.001 | 87.72 (17.08) | 86.95 (16.71) | <0.001 | 87.63 (16.83) | 86.97 (16.73) | 0.001 |
| CKD ≥3 (%) | 6,713 (3.5) | 777 (3.6) | 5,936 (3.5) | 0.281 | 605 (3.8) | 6,108 (3.5) | 0.051 | 430 (3.5) | 6,283 (3.5) | 0.888 | 269 (3.5) | 6,444 (3.5) | 1 |
| Hypertension (%) | 76,180 (38.0) | 8,516 (38.0) | 67,664 (38.0) | 0.892 | 6,331 (37.7) | 69,849 (38.0) | 0.434 | 4,846 (38.0) | 71,334 (38.0) | 0.911 | 3,062 (38.1) | 73,118 (38.0) | 0.759 |
| Pre-excitation syndrome (%) | 208 (0.1) | 20 (0.1) | 188 (0.1) | 0.548 | 16 (0.1) | 192 (0.1) | 0.819 | 10 (0.1) | 198 (0.1) | 0.44 | 10 (0.1) | 198 (0.1) | 0.677 |

Demographic and clinical characteristics of the overall study population, putative pathogenic variant carriers (G+), and those without any putative pathogenic variants in any CMP-associated, DCM-associated, HCM-associated, and ARVC-associated genes (G-). For continuous variables, standard deviation is shown in brackets.

ARVC, arrhythmogenic right ventricular cardiomyopathy; BP, blood pressure; DCM, dilated cardiomyopathy; CAD, coronary artery disease; CCD, cardiac conduction disease; CIED, cardiac implantable electronic device (including single- and dual-chamber permanent pacemaker, implantable cardioverter defibrillator, cardiac resynchronization therapy); CMP, inherited cardiomyopathy; CKD, chronic kidney disease; eGFR, estimated glomerular filtration rate; HCM, hypertrophic cardiomyopathy; PVC, premature ventricular complex.

**Table S3. The number of putative pathogenic variants in each of the ClinGen curated cardiomyopathy-associated genes.**

| Gene | n |
| --- | --- |
| *MYH6* | 1732 |
| *OBSCN* | 1712 |
| *>1 variant* | 1519 |
| *SCN5A* | 1498 |
| *MYH7* | 1247 |
| *TTN* | 1235 |
| *FLNC* | 1193 |
| *RYR2* | 1156 |
| *MYBPC3* | 500 |
| *CACNA1C* | 474 |
| *GAA* | 459 |
| *ABCC9* | 453 |
| *DSP* | 453 |
| *LAMA4* | 407 |
| *TNNI3K* | 401 |
| *DES* | 378 |
| *MYO6* | 371 |
| *PSEN2* | 369 |
| *LMNA* | 358 |
| *EYA4* | 294 |
| *ILK* | 250 |
| *ACTN2* | 249 |
| *MYOM1* | 249 |
| *DSG2* | 236 |
| *NEBL* | 205 |
| *LDB3* | 202 |
| *JUP* | 199 |
| *CSRP3* | 193 |
| *CTNNA3* | 189 |
| *MYPN* | 185 |
| *ALPK3* | 175 |
| *PKP2* | 175 |
| *SGCD* | 152 |
| *RBM20* | 147 |
| *TNNT2* | 143 |
| *DSC2* | 139 |
| *COX15* | 137 |
| *VCL* | 137 |
| *CDH2* | 134 |
| *TPM1* | 125 |
| *MYL3* | 117 |
| *TMEM43* | 113 |
| *MYL2* | 107 |
| *DTNA* | 102 |
| *PTPN11* | 101 |
| *NEXN* | 99 |
| *TGFB3* | 99 |
| *TBX20* | 98 |
| *TRIM63* | 95 |
| *CALR3* | 87 |
| *RAF1* | 87 |
| *CRYAB* | 85 |
| *JPH2* | 84 |
| *PLEKHM2* | 79 |
| *PRKAG2* | 74 |
| *SLC25A4* | 73 |
| *BAG3* | 72 |
| *MYLK2* | 72 |
| *PDLIM3* | 71 |
| *CAV3* | 62 |
| *GATAD1* | 62 |
| *TJP1* | 62 |
| *TNNI3* | 61 |
| *FXN* | 57 |
| *NKX2-5* | 57 |
| *PLN* | 55 |
| *TCAP* | 53 |
| *ACTC1* | 49 |
| *ANKRD1* | 47 |
| *TNNC1* | 43 |
| *FHL1* | 41 |
| *CTF1* | 38 |
| *MYOZ2* | 38 |
| *GLA* | 30 |
| *TTR* | 28 |
| *PRDM16* | 23 |
| *RIT1* | 22 |
| *LAMP2* | 15 |
| *KLF10* | 13 |

**Table S4. The number of putative pathogenic variants found in each of the ClinGen curated DCM-associated genes.**

| Gene | n |
| --- | --- |
| *MYH6* | 1790 |
| *OBSCN* | 1771 |
| *SCN5A* | 1516 |
| *TTN* | 1280 |
| *MYH7* | 1254 |
| *FLNC* | 1235 |
| *>1* variant | 848 |
| *MYBPC3* | 497 |
| *ABCC9* | 471 |
| *DSP* | 437 |
| *LAMA4* | 422 |
| *TNNI3K* | 415 |
| *PSEN2* | 384 |
| *DES* | 355 |
| *LMNA* | 354 |
| *EYA4* | 310 |
| *ILK* | 265 |
| *ACTN2* | 264 |
| *DSG2* | 241 |
| *NEBL* | 212 |
| *LDB3* | 210 |
| *CSRP3* | 199 |
| *MYPN* | 191 |
| *SGCD* | 155 |
| *RBM20* | 151 |
| *TNNT2* | 147 |
| *VCL* | 140 |
| *TPM1* | 131 |
| *MYL2* | 108 |
| *DTNA* | 105 |
| *NEXN* | 102 |
| *TBX20* | 101 |
| *JPH2* | 85 |
| *PLEKHM2* | 83 |
| *BAG3* | 73 |
| *TNNI3* | 65 |
| *GATAD1* | 62 |
| *NKX2-5* | 58 |
| *PLN* | 55 |
| *TCAP* | 54 |
| *ACTC1* | 49 |
| *ANKRD1* | 47 |
| *TNNC1* | 44 |
| *CTF1* | 39 |
| *PRDM16* | 23 |

Genes which are linked to more than one of DCM, HCM, and ARVC may have different numbers of putative pathogenic variants listed in Tables S3-5 due to different disease specific filtering allele frequencies.

ARVC, Arrhythmogenic right ventricular cardiomyopathy; DCM, Dilated cardiomyopathy; HCM, Hypertrophic cardiomyopathy.

**Table S5. The number of putative pathogenic variants found in each of the ClinGen curated HCM-associated genes.**

| Gene | n |
| --- | --- |
| *OBSCN* | 1261 |
| *RYR2* | 1222 |
| *TTN* | 1163 |
| *MYH6* | 1085 |
| *FLNC* | 970 |
| *MYH7* | 912 |
| *>1 variant* | 559 |
| *CACNA1C* | 499 |
| *GAA* | 485 |
| *MYO6* | 397 |
| *ABCC9* | 346 |
| *MYBPC3* | 288 |
| *MYOM1* | 263 |
| *DES* | 229 |
| *ACTN2* | 197 |
| *MYPN* | 187 |
| *ALPK3* | 181 |
| *LDB3* | 151 |
| *COX15* | 145 |
| *TNNT2* | 130 |
| *TPM1* | 117 |
| *PTPN11* | 108 |
| *CSRP3* | 100 |
| *TRIM63* | 100 |
| *RAF1* | 92 |
| *CRYAB* | 91 |
| *VCL* | 91 |
| *CALR3* | 90 |
| *NEXN* | 86 |
| *SLC25A4* | 84 |
| *JPH2* | 81 |
| *MYL2* | 80 |
| *PDLIM3* | 79 |
| *PRKAG2* | 77 |
| *MYLK2* | 75 |
| *BAG3* | 71 |
| *MYL3* | 70 |
| *CAV3* | 67 |
| *FXN* | 58 |
| *TCAP* | 55 |
| *TNNI3* | 52 |
| *TNNC1* | 45 |
| *FHL1* | 44 |
| *ANKRD1* | 43 |
| *MYOZ2* | 39 |
| *ACTC1* | 35 |
| *GLA* | 32 |
| *TTR* | 32 |
| *PLN* | 28 |
| *RIT1* | 22 |
| *LAMP2* | 17 |
| *KLF10* | 14 |

Genes which are linked to more than one of DCM, HCM, and ARVC may have different numbers of putative pathogenic variants listed in Tables S3-5 due to different disease specific filtering allele frequencies.

ARVC, Arrhythmogenic right ventricular cardiomyopathy; DCM, Dilated cardiomyopathy; HCM, Hypertrophic cardiomyopathy.

**Table S6. The number of putative pathogenic variants found in each of the ClinGen curated ARVC-associated genes.**

| Gene | n |
| --- | --- |
| *SCN5A* | 1620 |
| *MYH7* | 1363 |
| *TTN* | 1332 |
| *MYBPC3* | 539 |
| *DSP* | 495 |
| *DES* | 409 |
| *LMNA* | 384 |
| *DSG2* | 256 |
| *>1 variant* | 234 |
| *JUP* | 218 |
| *CTNNA3* | 210 |
| *PKP2* | 192 |
| *DSC2* | 149 |
| *CDH2* | 145 |
| *MYL3* | 131 |
| *TMEM43* | 120 |
| *TGFB3* | 110 |
| *TJP1* | 66 |
| *PLN* | 55 |

Genes which are linked to more than one of DCM, HCM, and ARVC may have different numbers of putative pathogenic variants listed in Tables S3-5 due to different disease specific filtering allele frequencies.

ARVC, Arrhythmogenic right ventricular cardiomyopathy; DCM, Dilated cardiomyopathy; HCM, Hypertrophic cardiomyopathy.

**Table S7**. **Cox proportional hazards regression comparing mortality for G+ to G-, stratified by ClinGen evidence level.**

|  | term | HR | p value |
| --- | --- | --- | --- |
| CMP | Limited | 1.09 (95% CI 1.00-1.18) | 0.040 |
|  | Moderate | 0.84 (95% CI 0.63-1.12) | 0.232 |
|  | Definitive/Strong | 1.08 (95% CI 1.00-1.16) | 0.039 |
| DCM | Limited | 1.08 (95% CI 0.99-1.17) | 0.079 |
|  | Moderate | 0.95 (95% CI 0.72-1.26) | 0.726 |
|  | Definitive/Strong | 1.10 (95% CI 1.00-1.20) | 0.044 |
| HCM | Limited | 1.14 (95% CI 1.04-1.26) | 0.006 |
|  | Moderate | 0.63 (95% CI 0.41-0.95) | 0.027 |
|  | Definitive/Strong | 1.06 (95% CI 0.96-1.18) | 0.248 |
| ARVC | Limited | 1.08 (95% CI 0.98-1.19) | 0.127 |
|  | Moderate | 1.11 (95% CI 0.78-1.57) | 0.557 |
|  | Definitive/Strong | 0.99 (95% CI 0.81-1.22) | 0.942 |

ARVC, Arrhythmogenic right ventricular cardiomyopathy; CMP, cardiomyopathy; DCM, Dilated cardiomyopathy; HCM, Hypertrophic cardiomyopathy; HR, hazard ratio; CI, confidence interval.

**Table S8**. **Cox proportional hazard regression comparing risk of developing a clinical cardiomyopathy for G+ to G-, stratified by ClinGen evidence level.**

|  | term | HR | p value |
| --- | --- | --- | --- |
| CMP | Limited | 1.334 (95% CI 0.956-1.863) | 0.09 |
|  | Moderate | 2.684 (95% CI 1.273-5.658) | 0.009 |
|  | Definitive/Strong | 3.245 (95% CI 2.63-4.004) | <0.0001 |
| DCM | Limited only | 1.179 (95% CI 0.79-1.761) | 0.412 |
|  | Limited | 2.942 (95% CI 1.574-5.5) | 0.0007 |
|  | Moderate | 2.844 (95% CI 1.349-5.995) | 0.006 |
|  | Definitive/Strong | 3.958 (95% CI 3.137-4.994) | <0.0001 |
| HCM | Limited only | 1.34 (95% CI 0.838-2.143) | 0.221 |
|  | Limited | 9.613 (95% CI 6.975-13.25) | <0.0001 |
|  | Moderate | 2.928 (95% CI 1.214-7.062) | 0.017 |
|  | Definitive/Strong | 2.256 (95% CI 1.63-3.121) | <0.0001 |
| ARVC | Limited only | 0.716 (95% CI 0.179-2.869) | 0.637 |
|  | Limited | 4.328 (95% CI 3.349-5.593) | <0.0001 |
|  | Moderate | 2.11 (95% CI 0.679-6.563) | 0.197 |
|  | Definitive/Strong | 2.531 (95% CI 1.394-4.596) | 0.002 |

The limited category is split into two groups depending on whether the gene has Definitive/Strong/Moderate evidence for another inherited cardiomyopathy (labelled “Limited”) or Limited/No evidence for another inherited cardiomyopathy (labelled “Limited only”).

ARVC, Arrhythmogenic right ventricular cardiomyopathy; CMP, cardiomyopathy; DCM, Dilated cardiomyopathy; HCM, Hypertrophic cardiomyopathy; HR, hazard ratio; CI, confidence interval.

**Table S9**. **Cox proportional hazards regression comparing composite outcomes** **for G+ to G-, stratified by ClinGen evidence level.**

|  | Term | HR | p value |
| --- | --- | --- | --- |
| CMP | Limited | 1.09 (95% CI 1.03-1.15) | 0.002 |
|  | Moderate | 0.97 (95% CI 0.80-1.18) | 0.787 |
|  | Definitive/Strong | 1.14 (95% CI 1.08-1.20) | <0.00001 |
| DCM | Limited | 1.09 (95% CI 1.03-1.16) | 0.003 |
|  | Moderate | 0.96 (95% CI 0.79-1.17) | 0.707 |
|  | Definitive/Strong | 1.20 (95% CI 1.12-1.27) | <0.00001 |
| HCM | Limited | 1.24 (95% CI 1.16-1.33) | <0.00001 |
|  | Moderate | 0.85 (95% CI 0.65-1.10) | 0.215 |
|  | Definitive/Strong | 1.05 (95% CI 0.98-1.13) | 0.182 |
| ARVC | Limited | 1.23 (95% CI 1.15-1.32) | <0.00001 |
|  | Moderate | 1.08 (95% CI 0.84-1.39) | 0.534 |
|  | Definitive/Strong | 1.03 (95% CI 0.89-1.19) | 0.727 |

The composite outcome includes: mortality, heart failure, stroke, atrial fibrillation, ventricular arrhythmias, and cardiac implantable electronic device insertion.

ARVC, Arrhythmogenic right ventricular cardiomyopathy; CMP, cardiomyopathy; DCM, Dilated cardiomyopathy; HCM, Hypertrophic cardiomyopathy; HR, hazard ratio; CI, confidence interval.

**Table S10**. **Cox proportional hazards regression comparing components of outcomes** **for G+ to G-, stratified by ClinGen evidence.**

|  | term | HR | p value |
| --- | --- | --- | --- |
| AF | CMP-G+ | 1.15 (95% CI 1.08-1.21) | 4.57E-06 |
|  | DCM-G+ | 1.18 (95% CI 1.11-1.26) | 5.58E-07 |
|  | HCM-G+ | 1.19 (95% CI 1.1-1.28) | 5.69E-06 |
|  | ARVC-G+ | 1.29 (95% CI 1.18-1.41) | 1.53E-08 |
| CIED | CMP-G+ | 1.15 (95% CI 1.04-1.28) | 0.006037 |
|  | DCM-G+ | 1.18 (95% CI 1.05-1.32) | 0.005538 |
|  | HCM-G+ | 1.12 (95% CI 0.98-1.28) | 0.098926 |
|  | ARVC-G+ | 1.19 (95% CI 1.02-1.4) | 0.029414 |
| Heart failure | CMP-G+ | 1.221 (95% CI 1.122-1.328) | 3.63E-06 |
|  | DCM-G+ | 1.288 (95% CI 1.173-1.414) | 1.13E-07 |
|  | HCM-G+ | 1.267 (95% CI 1.138-1.41) | 1.51E-05 |
|  | ARVC-G+ | 1.385 (95% CI 1.22-1.572) | 4.56E-07 |
| Cardiomyopathy | CMP-G+ | 2.377 (95% CI 1.98-2.853) | 1.46E-20 |
|  | DCM-G+ | 2.596 (95% CI 2.135-3.156) | 1.08E-21 |
|  | HCM-G+ | 2.788 (95% CI 2.257-3.443) | 1.75E-21 |
|  | ARVC-G+ | 3.435 (95% CI 2.722-4.334) | 2.46E-25 |
| Stroke | CMP-G+ | 0.99 (95% CI 0.89-1.11) | 0.892882 |
|  | DCM-G+ | 1 (95% CI 0.88-1.14) | 0.985935 |
|  | HCM-G+ | 1 (95% CI 0.86-1.15) | 0.947513 |
|  | ARVC-G+ | 1.03 (95% CI 0.87-1.23) | 0.727284 |
| Ventricular arrhythmia | CMP-G+ | 1.17 (95% CI 1.01-1.37) | 0.039665 |
|  | DCM-G+ | 1.19 (95% CI 1-1.41) | 0.050539 |
|  | HCM-G+ | 1.24 (95% CI 1.03-1.51) | 0.026374 |
|  | ARVC-G+ | 1.36 (95% CI 1.09-1.71) | 0.007355 |
| Premature ventricular complex | CMP-G+ | 1.127 (95% CI 0.861-1.477) | 0.384 |
|  | DCM-G+ | 1.207 (95% CI 0.896-1.625) | 0.216 |
|  | HCM-G+ | 1.343 (95% CI 0.972-1.855) | 0.0742 |
|  | ARVC-G+ | 1.53 (95% CI 1.051-2.226) | 0.0263 |

Results are adjusted for sex.

ARVC, Arrhythmogenic right ventricular cardiomyopathy; CMP, cardiomyopathy; CIED, cardiac implantable electronic device; DCM, Dilated cardiomyopathy; HCM, Hypertrophic cardiomyopathy; HR, hazard ratio; CI, confidence interval.

**STROBE Statement**—Checklist of items that should be included in reports of ***cohort studies***

|  | Item No | Recommendation | Page Number |
| --- | --- | --- | --- |
| **Title and abstract** | 1 | (*a*) Indicate the study’s design with a commonly used term in the title or the abstract | 3 |
|  |  | (*b*) Provide in the abstract an informative and balanced summary of what was done and what was found | 3 |
| Introduction | | |  |
| Background/rationale | 2 | Explain the scientific background and rationale for the investigation being reported | 6 |
| Objectives | 3 | State specific objectives, including any prespecified hypotheses | 6 |
| Methods | | |  |
| Study design | 4 | Present key elements of study design early in the paper | 7-9 |
| Setting | 5 | Describe the setting, locations, and relevant dates, including periods of recruitment, exposure, follow-up, and data collection | 7-9 |
| Participants | 6 | (*a*) Give the eligibility criteria, and the sources and methods of selection of participants. Describe methods of follow-up | 7-9 |
|  |  | (*b*) For matched studies, give matching criteria and number of exposed and unexposed | NA |
| Variables | 7 | Clearly define all outcomes, exposures, predictors, potential confounders, and effect modifiers. Give diagnostic criteria, if applicable | 7-9, Table 1 |
| Data sources/ measurement | 8* | For each variable of interest, give sources of data and details of methods of assessment (measurement). Describe comparability of assessment methods if there is more than one group | 7-9 |
| Bias | 9 | Describe any efforts to address potential sources of bias | 7-9 |
| Study size | 10 | Explain how the study size was arrived at | 7-9 |
| Quantitative variables | 11 | Explain how quantitative variables were handled in the analyses. If applicable, describe which groupings were chosen and why | 7-9 |
| Statistical methods | 12 | (*a*) Describe all statistical methods, including those used to control for confounding | 9, supplementary methods |
|  |  | (*b*) Describe any methods used to examine subgroups and interactions | 7-9 |
|  |  | (*c*) Explain how missing data were addressed | NA |
|  |  | (*d*) If applicable, explain how loss to follow-up was addressed | NA |
|  |  | (*e*) Describe any sensitivity analyses | NA |
| Results | | |  |
| Participants | 13* | (a) Report numbers of individuals at each stage of study—eg numbers potentially eligible, examined for eligibility, confirmed eligible, included in the study, completing follow-up, and analysed | 10, Table 1 |
|  |  | (b) Give reasons for non-participation at each stage | NA |
|  |  | (c) Consider use of a flow diagram | NA |
| Descriptive data | 14* | (a) Give characteristics of study participants (eg demographic, clinical, social) and information on exposures and potential confounders | 10, Table 1 |
|  |  | (b) Indicate number of participants with missing data for each variable of interest | NA |
|  |  | (c) Summarise follow-up time (eg, average and total amount) | NA |
| Outcome data | 15* | Report numbers of outcome events or summary measures over time | Figure 1-5 |
| Main results | 16 | (*a*) Give unadjusted estimates and, if applicable, confounder-adjusted estimates and their precision (eg, 95% confidence interval). Make clear which confounders were adjusted for and why they were included | 10-12, Figure 1-5 |
|  |  | (*b*) Report category boundaries when continuous variables were categorized | NA |
|  |  | (*c*) If relevant, consider translating estimates of relative risk into absolute risk for a meaningful time period | NA |
| Other analyses | 17 | Report other analyses done—eg analyses of subgroups and interactions, and sensitivity analyses | NA |
| Discussion | | |  |
| Key results | 18 | Summarise key results with reference to study objectives | 13 |
| Limitations | 19 | Discuss limitations of the study, taking into account sources of potential bias or imprecision. Discuss both direction and magnitude of any potential bias | 17 |
| Interpretation | 20 | Give a cautious overall interpretation of results considering objectives, limitations, multiplicity of analyses, results from similar studies, and other relevant evidence | 14-16 |
| Generalisability | 21 | Discuss the generalisability (external validity) of the study results | 13-16 |
| Other information | | |  |
| Funding | 22 | Give the source of funding and the role of the funders for the present study and, if applicable, for the original study on which the present article is based | 2 |

*Give information separately for exposed and unexposed groups.

**Figure S1. Kaplan-Meier plots comparing all-cause mortality in individuals with vs without putative pathogenic variants in cardiomyopathy genes.**

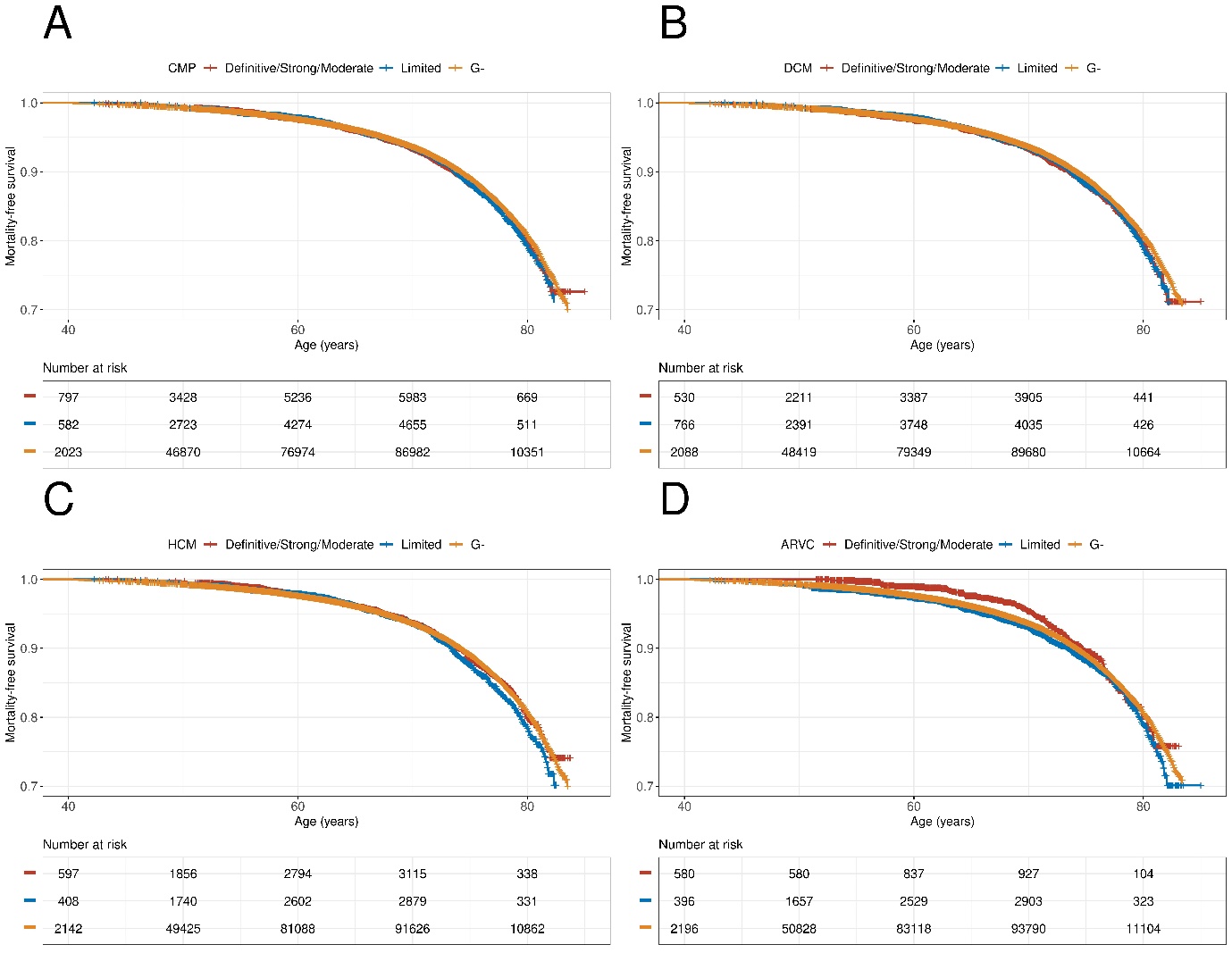

The curves show the mortality in individuals with putative pathogenic variants (PuPV) all cardiomyopathy-associated genes (A), DCM-associated genes (B), HCM-associated genes (C), and ARVC-associated gene (D) versus those without (G-) separated by ClinGen level of evidence. Hazard ratios are displayed, calculated using Cox proportional hazard regression, correct for sex and using age as timescale.

ARVC, Arrhythmogenic right ventricular cardiomyopathy; CMP, cardiomyopathy; DCM, Dilated cardiomyopathy; HCM, Hypertrophic cardiomyopathy.

**Figure S2. Kaplan-Meier plots comparing composite outcomes in individuals with vs without putative pathogenic variants in cardiomyopathy genes.**

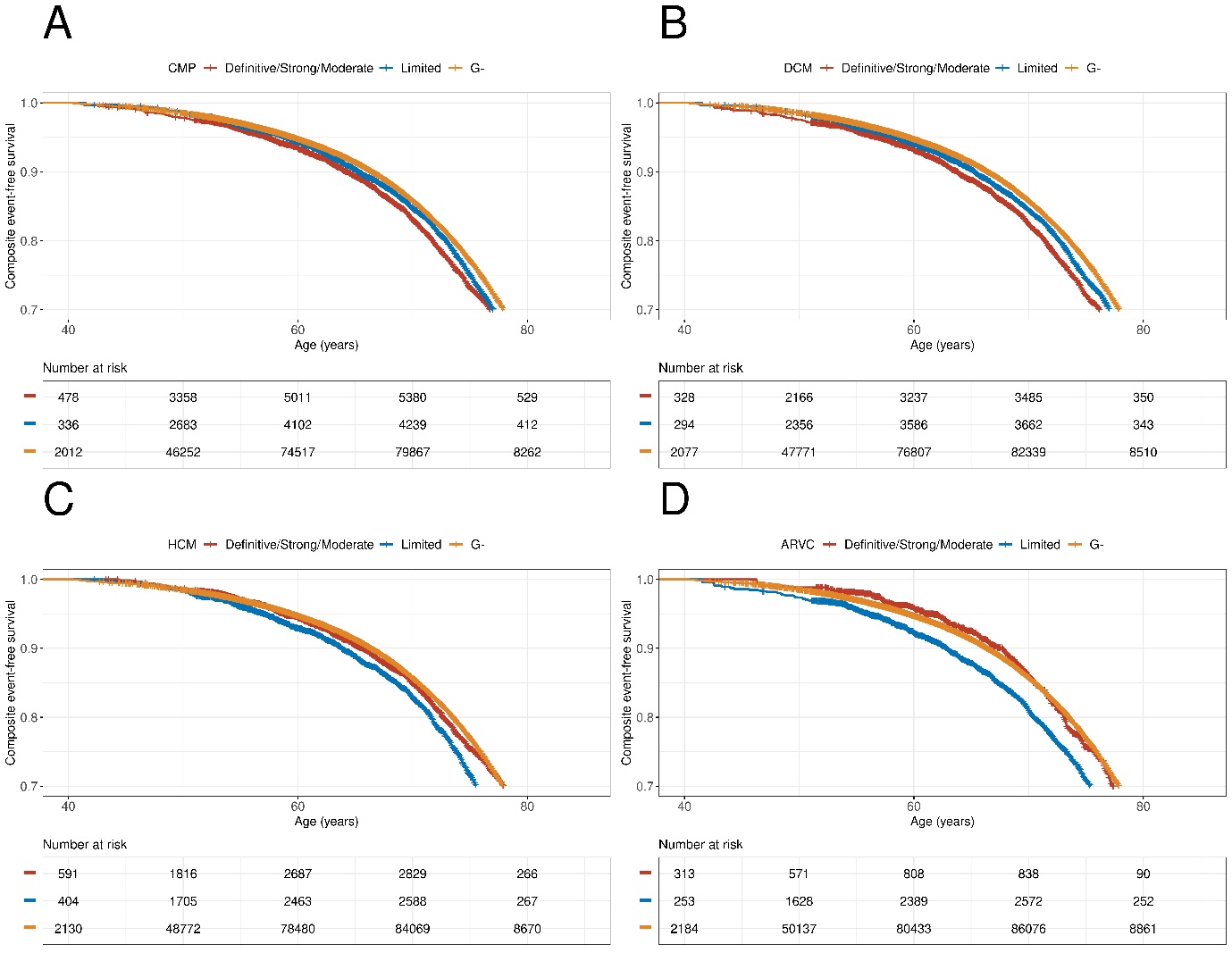

The composite outcomes include: mortality, heart failure, stroke, atrial fibrillation, ventricular arrhythmias, cardiac implantable electronic device insertion.

(A), DCM-associated genes (B), HCM-associated genes (C), and ARVC-associated gene (D) versus those without (G-) split by ClinGen level of evidence. Hazard ratios are displayed, calculated using Cox proportional hazard regression, corrected for sex and using age as timescale.

ARVC, Arrhythmogenic right ventricular cardiomyopathy; CMP, cardiomyopathy; DCM, Dilated cardiomyopathy; HCM, Hypertrophic cardiomyopathy.

**Figure S3. Kaplan-Meier plots comparing mortality in individuals with vs without putative pathogenic variants in HRS-defined high-risk cardiomyopathy genes and actionably cardiomyopathy genes, as defined by the ACMG.**

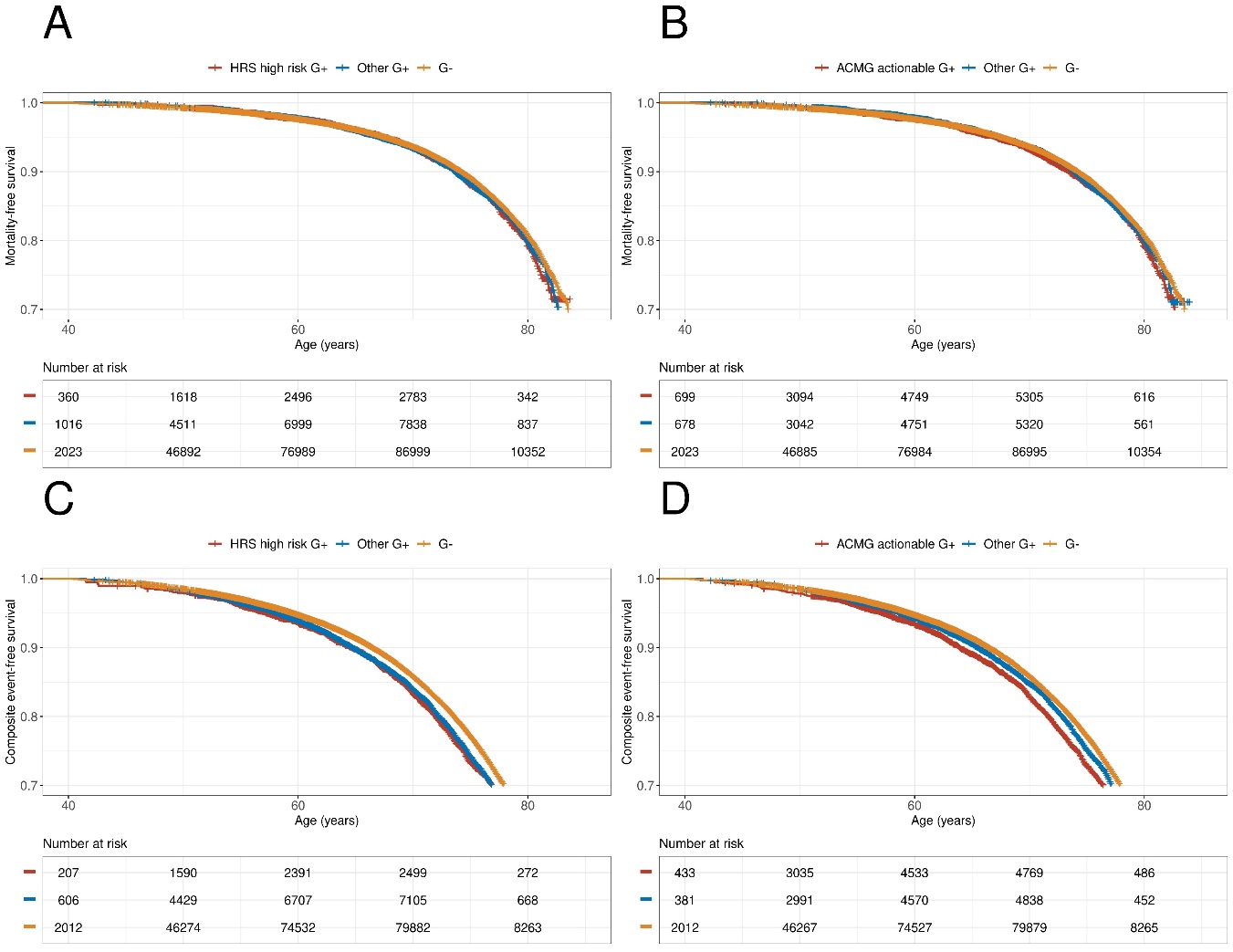

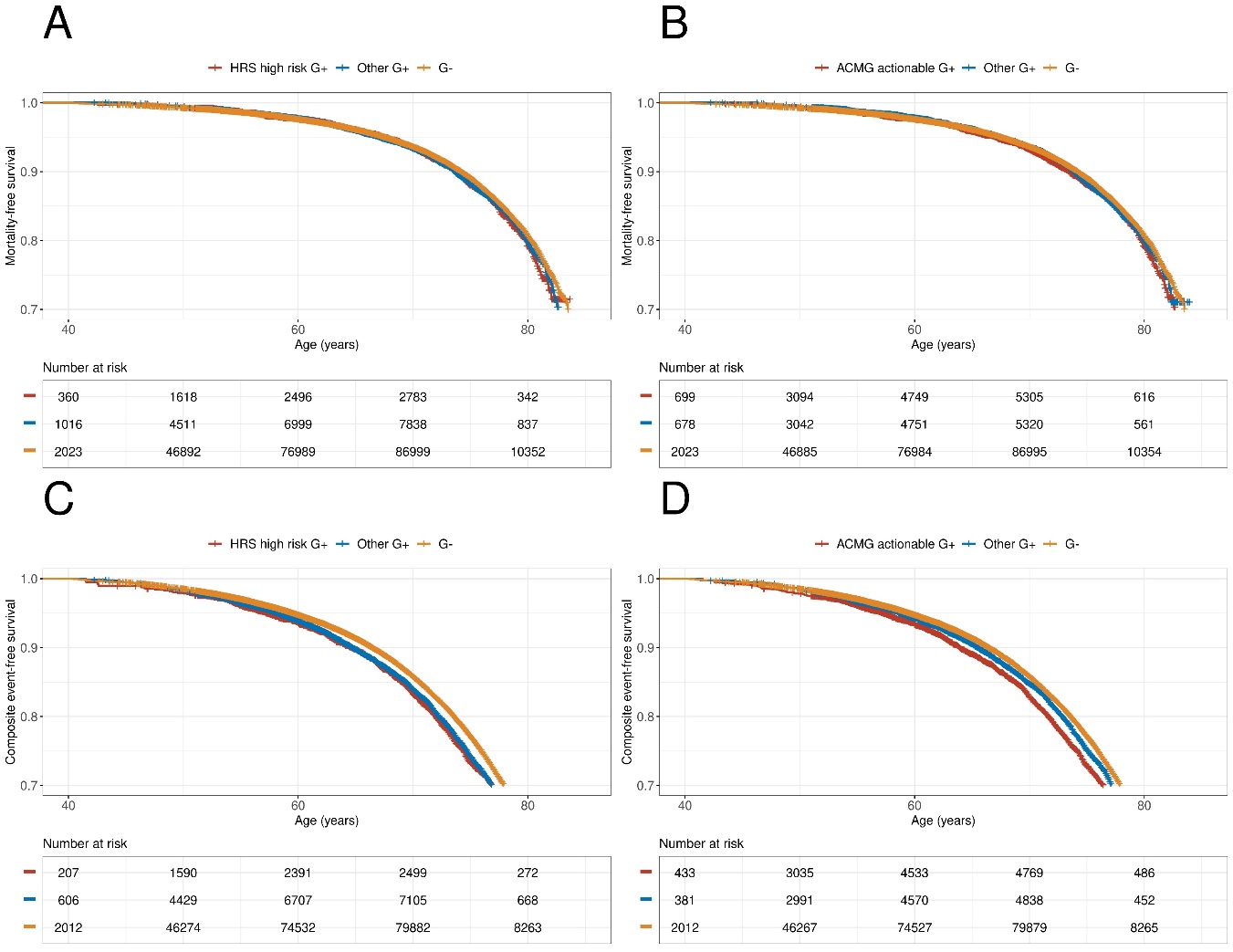

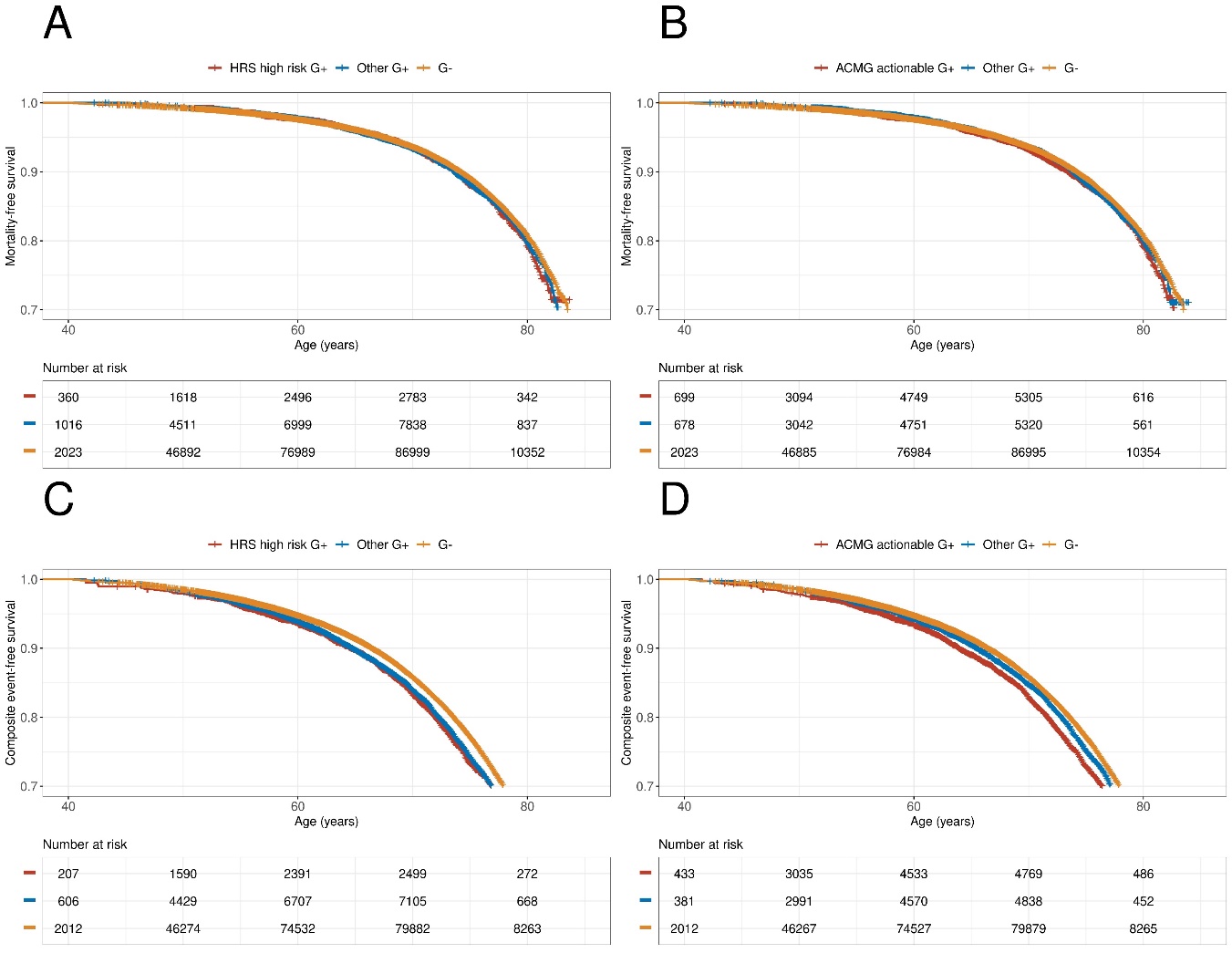

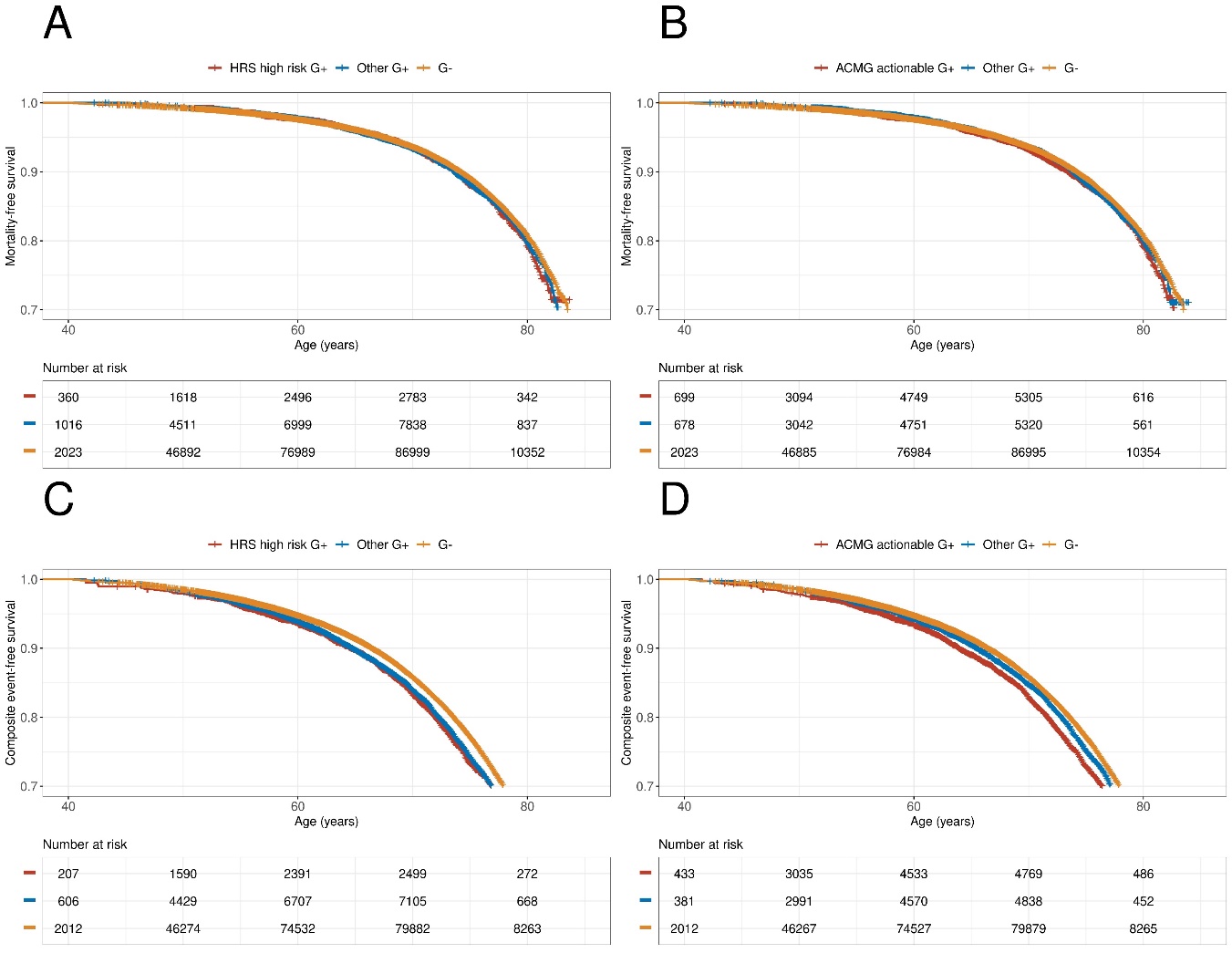

A

B

C

D

The composite outcomes include: mortality, heart failure, stroke, atrial fibrillation, ventricular arrhythmias, and cardiac implantable electronic device insertion.

Panels (A) and (B) compare individuals with putative pathogenic variants in HRS high risk genes vs any other cardiomyopathy gene vs those without any variants (G-)

(A) all-cause mortality and (B) the composite outcomes.

Panels (C) and (D) compare individuals with putative pathogenic variants in an ACMG actionable genes vs any other cardiomyopathy gene vs (G-)

(C) all-cause mortality and (D) the composite outcomes.

ARVC, Arrhythmogenic right ventricular cardiomyopathy; CMP, cardiomyopathy; DCM, Dilated cardiomyopathy; HCM, Hypertrophic cardiomyopathy.
